# Computational Analysis of Treatment Resistant Cancer Cells

**DOI:** 10.1101/2024.08.29.24312813

**Authors:** Alexandre Matov

## Abstract

**Introduction:** Prostate cancer (PC), which is a disease driven by the activity of the androgen receptor (AR), is the most commonly diagnosed malignancy and despite advances in diagnostic and treatment strategies, PC is the second most common cause of cancer mortality in men (Bray et al., 2018). Taxane-based chemotherapy is the only chemotherapy that prolongs survival in metastatic PC patients (Petrylak et al., 2004; Tannock et al., 2004). At the cellular level, taxanes bind to and stabilize microtubules (MTs) inhibiting all MT-dependent intracellular pathways. MTs are highly dynamic polymers that stochastically switch between phases of growth, shrinkage, and pause (Jordan and Wilson, 2004). Altered MT dynamics endow cancer cells with both survival and migratory advantages (Mitchison, 2012). Taxanes inhibit MT dynamics and alter the spatial organization of the MT network, thereby inhibiting intracellular trafficking of molecular cargo critical for tumor survival. In PC specifically, taxanes inhibit transcriptional activity downstream of MT stabilization (Thadani-Mulero et al., 2012) and AR nuclear accumulation (Darshan et al., 2011; Zhu et al., 2010).

**Methods:** Different tubulin inhibitors, even from within the same structural class as the taxanes, affect distinct parameters of MT dynamics (Jordan and Wilson, 2004), yet the selection of taxane for chemotherapy is not based on the particular patterns of dynamic behavior of the MT cytoskeleton in individual patients. We envisage that systematic characterization using quantitative analysis of MT dynamics in PC patient cells expressing clinically relevant protein isoforms (Matov and Bacconi, 2024; Thoma et al., 2010), before and after treatment with each of the taxanes, will allow us to identify criteria for the selection of the most suitable drug combination at the onset of treatment.

**Results:** We link MT dynamics in the presence of AR variants and sensitivity/resistance to taxanes and connect fundamental research with clinically relevant concepts to elucidate cellular mechanisms of clinical response to taxanes and, thus, advance the customization of therapy. Our computational live-cell analysis addresses questions in the context of the inherent differences in MT homeostasis as a function of AR content in PC cells, the specific parameters of MT dynamics each of the taxanes affects, and how can this information be used to match endogenous patterns of MT dynamics with drug-modulated MT behavior.

**Conclusions:** We investigate whether the sensitivity to taxanes, evaluated by computational analysis of MTs, can be linked to gene expression correlated with AR and its variants, and whether the resistance to taxanes can be linked to the presence of a specific AR splice variant, and can we identify which of the taxanes will be most effective based on the endogenous patterns of MT dynamics.

## INTRODUCTION

Though treatment options for metastatic castration-resistant prostate cancer (mCRPC) have expanded over the past decade, highly proliferative phenotypes frequently emerge at the time of progression on androgen signaling inhibitors. Though initially effective in reducing tumor burden for some patients, resistance to systemic therapy is universal and approximately one-third of tumors are primarily refractory to this treatment approach. This is clinically highly significant as mCRPC refractory to tubulin inhibitors is uniformly fatal within 12-18 months (Francini et al., 2019). Currently, mCRPC chemotherapy is limited to the FDA-approved docetaxel and cabazitaxel and could be potentially impactful. However, even for the FDA-approved taxanes, there is currently no good mechanistic understanding of drug action and, for instance, docetaxel is always used as a first line therapy without having to examine patient cells *ex vivo* for susceptibility (Antonarakis et al., 2017). For patients with progressive mCRPC that was refractory to AR pathway inhibitors and had received or been deemed ineligible for taxane chemotherapy, there are not many treatment options available. For patients with mutated BRCA1/2, treatment with olaparib offers a benefit (De Santis et al., 2024) and novel therapies are needed to extend survival.

AR wild type (wt) and AR splice variants differentially associate with MT polymers and MT dynamics regulate AR signaling in PC (Darshan et al., 2011; Zhu et al., 2010). AR binds MTs and, upon stimulation, traffics towards the nucleus and activates gene transcription (Thadani-Mulero et al., 2012). It is unlikely AR, with size of <0.1 μm, is transported toward MTs minus tips by cytoplasmic dynein, which traffics mRNAs (about five times larger in size than AR) and much larger molecular cargos, but rather via diffusion. In addition to AR-wt, AR splice variants are co-expressed in metastatic PC patients, such as ARv567 and AR-V7, which arise following castration (Guo et al., 2009; Hu et al., 2009; Sun et al., 2010; Watson et al., 2010). These variants, which lack the ligand-binding domain (but have the DNA binding and transactivation domains) are insensitive to androgen deprivation therapy (ADT), and are constitutively active in the nucleus, which allows for continuous AR transcriptional activity. The ARv567 variant was shown to be present in 59% of patients with metastatic PC and to arise in response to abiraterone (Mostaghel et al., 2011). A MT co-sedimentation assay using cells transfected with the two clinically relevant AR variants, ARv567 and ARv7, showed that the majority of ARv567 is associated with MTs while only 40% of ARv7 co-fractionated with MT polymers (Thadani-Mulero et al., 2014). Further, paclitaxel treatment inhibited ARv567 nuclear accumulation, while it had no effect on variant ARv7 (Thadani-Mulero et al., 2014). Importantly, the lack of ARv7 inhibition of nuclear accumulation is correlated with docetaxel resistance *in vivo* using a LuCAP xenograft model of PC (Nguyen et al., 2017). Furthermore, these *in vivo* data showed that the ARv567 variant is very sensitive to docetaxel treatment and the high level of sensitivity to docetaxel remains persistent for 26 weeks of treatment (Nguyen et al., 2017). Taken together, these data indicate a dependency of AR’s transcriptional activity on the dynamics of the MT cytoskeleton.

## RESULTS

### COMPUTATIONAL ANALYSIS OF MT DYNAMICS

Our computational approach for the analysis of MT dynamics in living cells is based on the automated tracking of the motion of MT growing (or plus) tips in high-resolution confocal microscopy image sequences using a fluorescently tagged MT plus-end marker EB1 or EB3 (end-binding protein 1 or 3) (Matov et al., 2010). EB1 and EB3 recognize conformational changes and bind the MT plus tips during polymerization (growth) (Slep and Vale, 2007). When EB1 (or EB3) are EGPF-labeled, the EB1ΔC-2xEGFP dimers at the MT plus tip form a fluorescent comet, which serves as a molecular marker for the direct visualization of MT polymerization dynamics (Akhmanova and Steinmetz, 2008). Our image analysis algorithm tracks thousands of EB1 comets visible in an image time-lapse sequence allowing the detection of spatial patterns of MT dynamics and introduces spatiotemporal clustering of EB1ΔC-2xEGFP growth tracks to infer MT behaviors during phases of pause and shortening. EB1 comet detection is accomplished by band-pass filtering implemented as a Difference of Gaussians (Wilson and Giese, 1977) operator (for comparisons with a Laplacian of Gaussian and the Harris operator, see (Lindeberg, 2014)), which enhances image features with the expected shape and size of an EB1 comet, while suppressing higher-frequency imaging noise and lower-frequency structures representing larger than the EB1 comets aggregates of non-specific fluorescently labeled proteins. We compute the sum of the squared intensity differences between each segmented EB1 comet and the average comet profile to exclude the few detections with unusual shapes due to labeling artifacts via an unimodal histogram thresholding (Rosin, 2001). Our EB1 comet detection algorithm is robust against variations in comet lengths and brightness, both within videos and between videos. The feature detector delivers the position of each comet in a time-frame as well as the eccentricity and angular orientation of the comets. The latter is used as a directional cue in the subsequent motion tracking of comets from one time-frame to the next.

The cost of linking comets is defined in statistical terms as the Mahalanobis distance between projected and extracted comets (Mahalanobis, 1936). Detected EB1 comets are then tracked using a linear Kalman filter (Swerling, 1959) based algorithm (Yang et al., 2005). The EB1 comets serve as a molecular marker for the direct visualization of MT polymerization dynamics only and we introduced spatiotemporal clustering of EB1ΔC-2xEGFP growth tracks to infer MT behaviors during the phases of pause and shortening. Clustering of collinear MT growth tracks that likely belong to the same MT is performed by solving the bipartite graph as a linear assignment problem (LAP) via the Hungarian algorithm (Kuhn, 1955). It identifies the globally optimal positional correspondences between the endpoints of EB1 tracks and the start points of new tracks, and every potential assignment is characterized by a cost of linking (Matov et al., 2010). Specifically, a pairing between an EB1 track termination and a track initiation is considered only if these two events are separated by less than 15 seconds or another use-specified time-lag. Furthermore, the track initiation must fall within either one of two cone-shaped, geometrical search regions. The choice of the forward cone with an opening half-angle of 60° is motivated by the observation that during pauses or out of focus movements, MTs sometimes undergo significant lateral displacements and directional changes, especially near the cell edge. In contrast, the backward cone has an opening half-angle of 10° only, motivated by the observation that a MT catastrophe generally follows the recent MT growth track until a MT rescue or the complete depolymerization of the MT. By construction, this track assignment is spatially and temporally global, which warrants a high level of robustness.

### CYTOSKELETON REGULATION IN PC

#### MT dynamics in M12 isogenic PC cell lines expressing GFP-tagged wt- or variant-AR

We used cells from an isogenic PC cell line series of the M12 cells expressing GFP-tagged wt-or variant-AR and measured MT polymerization rates in three of these M12 series - parental cells with no endogenous AR, AR-wt cells, and ARv7 variant cells - which simultaneously express EB1ΔC-2xEGFP. In metastatic PC patients, the treatment of tumors which express the ARv7 variant correlates with clinical taxane resistance (Hörnberg et al., 2011). Also, increased MT dynamicity has been correlated with taxane resistance (Gonçalves et al., 2001). Figure 1 shows the MT growth tracks (EB1 comet tracks) overlaid on raw image data from M12 cells stably expressing AR-wt (Fig. 1A) or ARv7 (Fig. 1B). In cells expressing the AR-wt, in comparison to parental cells not expressing AR, MT dynamicity did not change drastically (e.g., the average speed of EB1 comets was reduced to 16.2 μm/min (Fig. 1C) from 17.3 μm/min (Fig. S1) in the AR-null cells). However, when we expressed the ARv7 variant, EB1 speeds increased with over 40% to an average of 24.6 μm/min (Fig. 1D). In M12 AR-wt cells, AR is localized both in the cytoplasm and the nucleus (Fig. 1A, the overall “haze” visible in the background of the images is a result of the fluorescent AR). In M12 ARv7 cells, AR localization was predominantly nuclear (Fig. 1B, see the brighter nuclear areas), in agreement with published data (Guo et al., 2009). Additionally, in cells expressing AR-wt, MT growth rates slow down considerably as MTs approach the cell edge (Fig. 1A), while no such slowdown was observed in M12 ARv7 cells (Fig. 1B). We also clearly observed differential spatial organizations of the MT cytoskeleton, with M12 ARv7 cells’ EB1 tracks rarely overlapping with the nuclear areas. Together, the results indicate differential regulation in taxane sensitive and taxane resistant cells.

**Figure 1.**
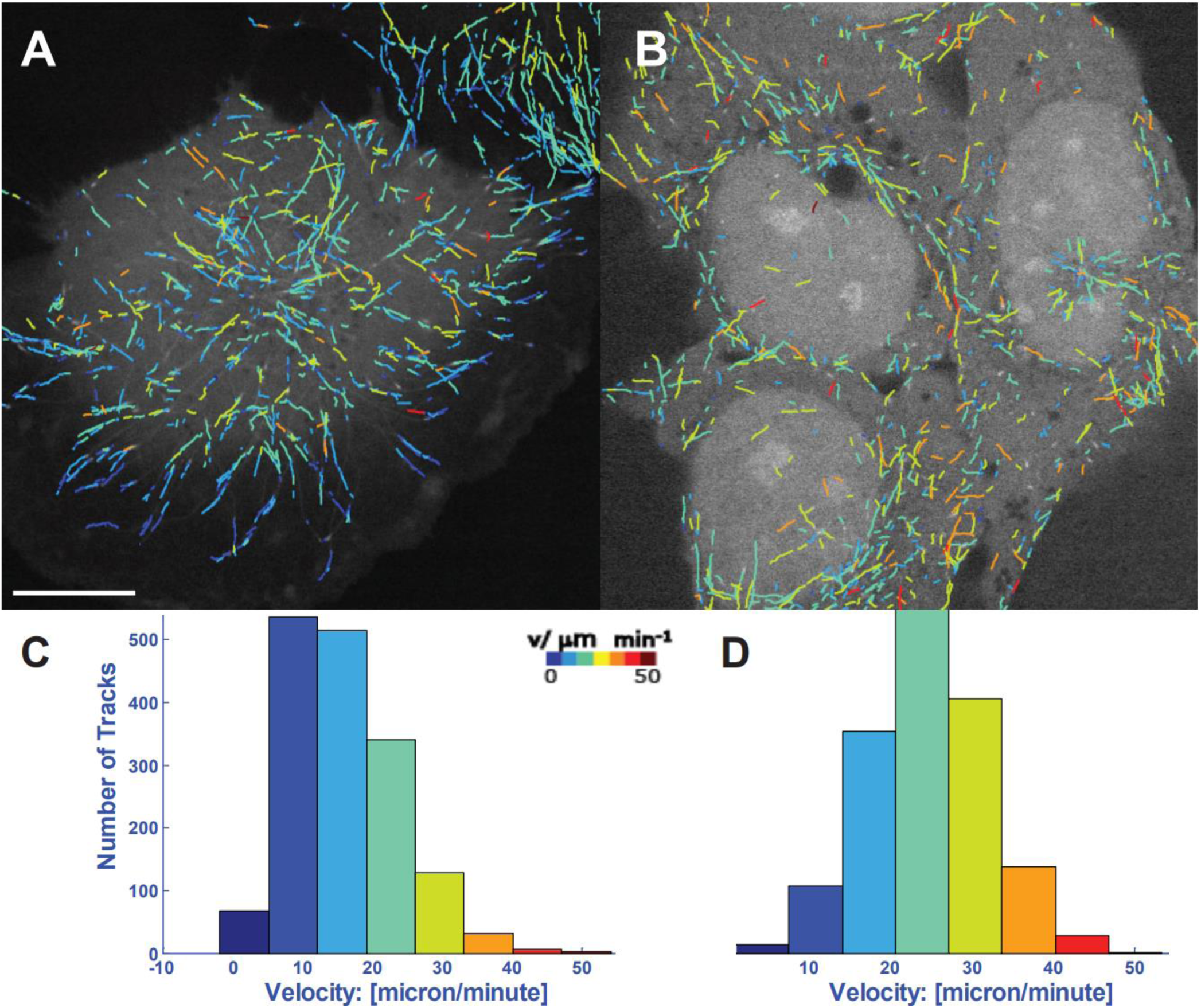
Tracking of EB1 comets in M12 isogenic PC cell lines expressing GFP-tagged wt- or variant-AR. MT tips and AR are labeled with GFP and imaged for a minute (with an acquisition rate of two images per second). EB1 comets are computationally tracked (Yang et al., 2005). The color-coding represents EB1 speeds and colder colors correspond to lower speeds, and warmer colors correspond to faster speeds. Scale bar equals 5 µm. (A) MT growth tracks for PC cells expressing the wild type AR variant. The median speed was about 15 µm with a marked slowdown at the edge, where there was no AR. (B) MT growth tracks for cells expressing ARv7 variant which is resistant to paclitaxel treatment. The median speeds were about 24 um/min. The lower panels show the corresponding EB1 comet velocity histograms. Histograms of growth velocities in µm/min are shown on (C) for AR wild type and (D) for the ARv7 variant.

#### Patient-derived organoids

Localized PC can be cured through surgical intervention. However, for the patients who are not good candidates for this intervention, e.g., those with high risk localized tumors or with locally advanced or metastatic disease, the question of whether therapy with tubulin inhibitors would be beneficial is of utmost importance. Clinically, the use of tubulin inhibitors to treat advanced PC is expected to significantly increase based on the results of the CHAARTED (Sweeney et al., 2015) and STAMPEDE (Vale et al., 2016) studies, which showed about 14 and 22 months survival benefit, respectively, based on combination of ADT and taxane therapy. Similarly, the STAMPEDE trial showed the benefit of docetaxel for early treatment of locally advanced tumors. Approximately one third of tumors are primarily refractory to this treatment approach, yet markers predicting drug efficacy are not available and the mechanisms of resistance are not understood. Therefore, there is an unmet clinical need to match individual tumors to a tubulin inhibitor most likely to be efficacious (Matov, 2024d). We propose to identify such matches based on MT dynamics signatures in metastatic PC organoids (Gao et al., 2014) and derived organoids from primary (UCSF-PR), micro-metastatic (UCSF-LN), and distant metastasis (UCSF-PCa) tumor tissues from 16 PC patients.

We dissociated prostate tissues (Fig. 2) according to (Goldstein et al., 2011) and cultured organoids according to (Drost et al., 2016; Gao et al., 2014). The main hurdle in culturing of primary prostate tumors has been the lack of markers to select against normal tissue cells (Drost et al., 2016). In colorectal cancer organoids selection against normal cells is done by removing R-spondin1, an effector of Wnt signaling, from the medium (van de Wetering et al., 2015). However, there is no such molecular marker in primary PC. Because we were aware of the issue of mixed tissue in primary tumors, we initially tried to minimize the amount of the sample by using a specialized twin-needle (Fig. 3), a gift from Paul Wegener, Epitome Pharmaceuticals, to increase the likelihood of capturing the tumor tissue only as well as to have a matching tissue for histology. However, we found tissue with Gleason Score 5, 4, 3 as well as normal tissue within less than 1 mg sample (Fig. 3A).

**Figure 2.**
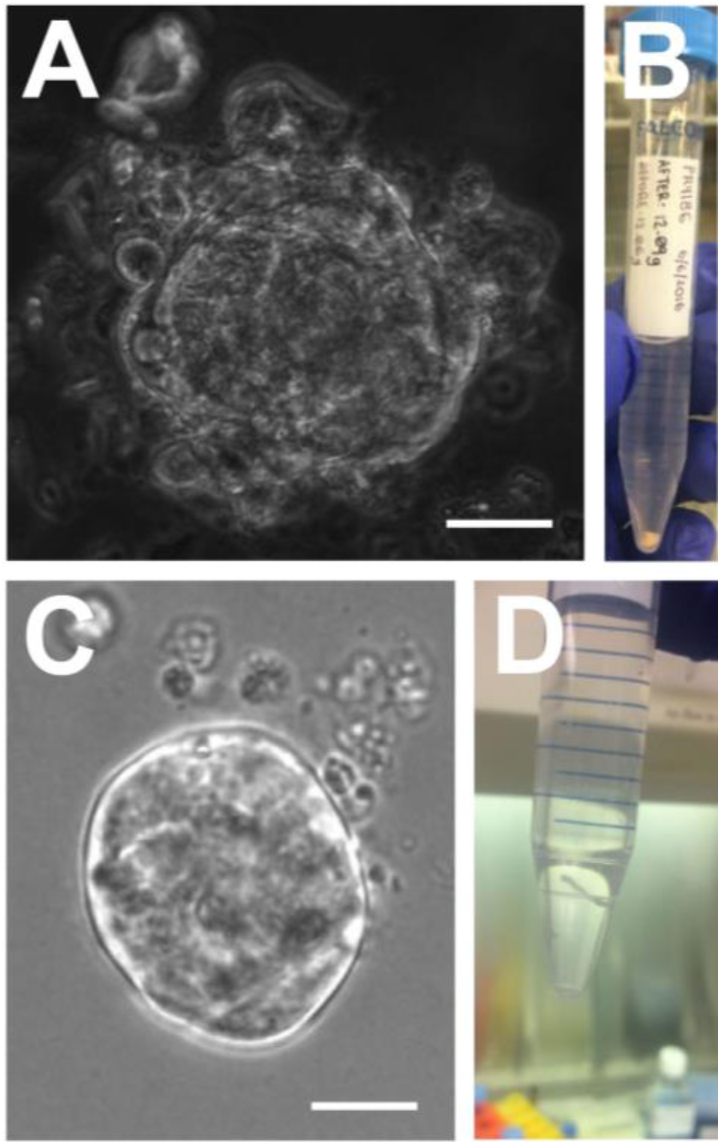
Phase contrast and transmitted light live-cell images of novel organoids. (A) UCSF-PR6 (primary tumor) on Day 29 after plating of single tumor cells. Scale bar equals 50 μm. (B) Tissue (3 mg) dissociated to derive UCSF-PR6 from 3 mm punch core biopsy. (C) UCSF-PCa1 (metastatic tumor, visceral tissue (liver)) organoid on Day 14 after plating of single tumor cells. Scale bar equals 30 μm. (D) Tissue (1 mg) dissociated to derive UCSF-PCa1 from needle biopsy.

**Figure 3.**
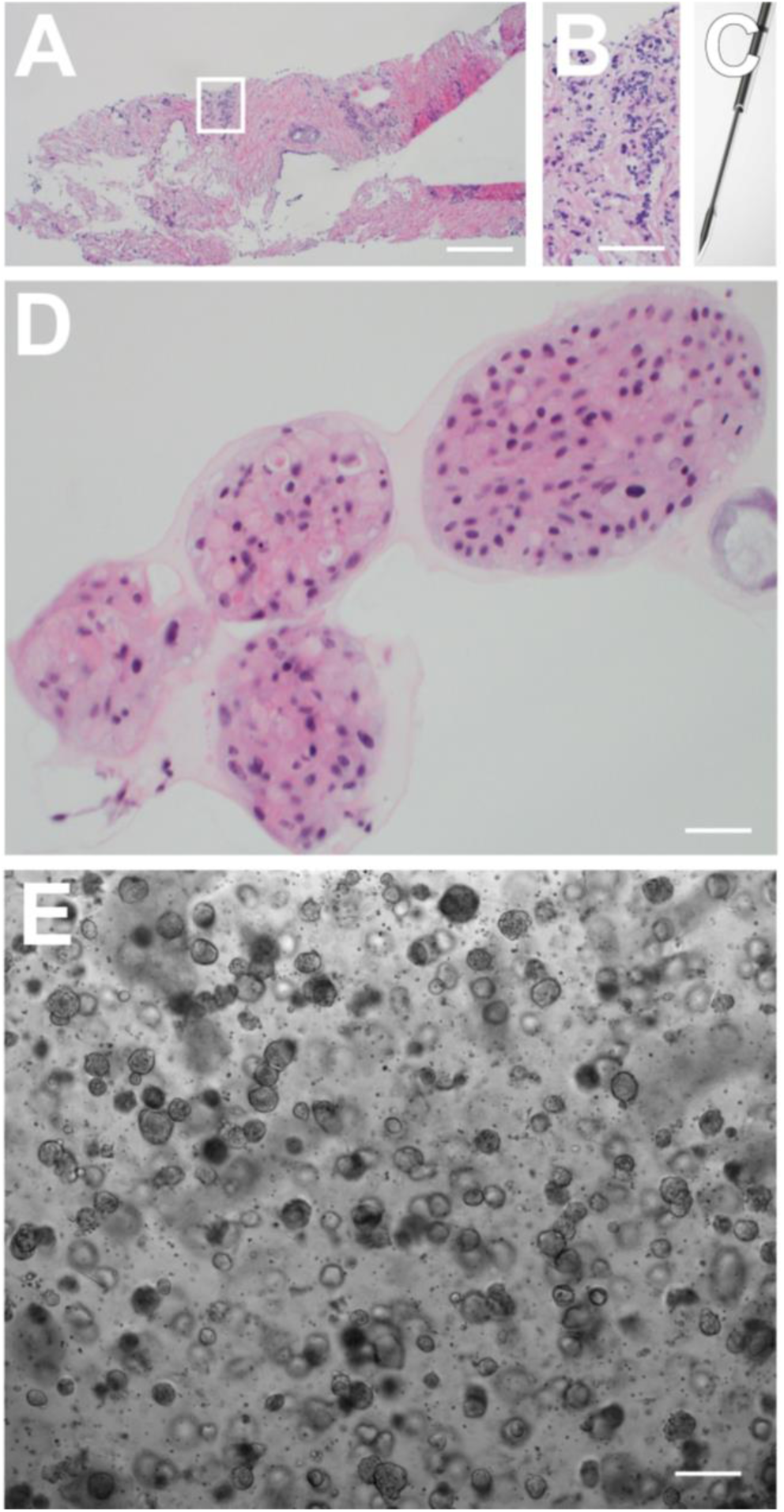
PC tissue histology, organoid culture and histology. (A) H&E of the tissue sample for UCSF-PR5 obtained with Twin-needle biopsy, Gleason Score 5+3+4; in the white rectangle is area of high grade tumor (Gleason Score 5). Scale bar equals 100 μm. (B) Enlarged zone of tumor pattern with Gleason Score 5 from the white rectangle in (A). Scale bar equals 40 μm. (C) Twin-needle provides matching tissue pairs, one for organoid culture and one for histology (shown on (A)). (D) Pathology assessment determines benign organoid culture by histology of UCSF-PR4 derived from 2 mg punch core biopsy tissue with Gleason Score 4+3 and 10% tumor density. Scale bar equals 20 μm. (E) Transmitted light image of small size round organoids, from UCSF-PR7 on Day 10 after seeding of single patient cells. Scale bar equals 200 μm.

#### Y-27632 ROCK inhibitor

Next, we made an effort to optimize the organoid medium composition to select for prostate tumor cells. Because normal and tumor tissue have distinctly different requirements for matrix-dependent signaling (see Fig. 4, increased E-cadherin expression in tumor tissue compared to the benign tissue used for organoid UCSF-PR2), correct modulation of contractility is important for successful tumor organoid establishment. Y-27632 is an inhibitor of Rho-kinase, which inhibits myosin light chain phosphorylation and as a result myosin activity (Matov, 2024f; Matov, 2025b; Matov and Bacconi, 2024; Narumiya et al., 2000). The inhibition of acto-myosin contractility is advantageous for establishing 3D organoid cultures (Sachs and Clevers, 2014), because it modulates the mechanical interactions with the extracellular matrix provided by the Matrigel. Y-27632 is added during phases of establishment of prostate organoids from single cells (Drost et al., 2016) to prevent apoptosis during the procedure, which includes long-term suspension and soft Matrigel culture. By seeding organoids in different wells with different levels of the drug in the medium, we observed that cells from benign tissues are considerably much more dependent on Y-27632 (see the effects of 70 µM of the myosin II inhibitor blebbistatin – which accurately replicates all of the effects of Y-27632 – on the contractility of the actin cytoskeleton in (Matov, 2024f)) ROCK inhibitor to form organoids. Also, we have found empirically that cells derived from high grade prostate tumors are resilient (Green, 2024) to apoptosis and have consequently been able to temporarily select tumor cells by reducing the concentration of Y-27632 ROCK inhibitor 4-fold (to 2.5 µM) in comparison to the protocol for normal prostate tissue organoids (Drost et al., 2016).

**Figure 4.**
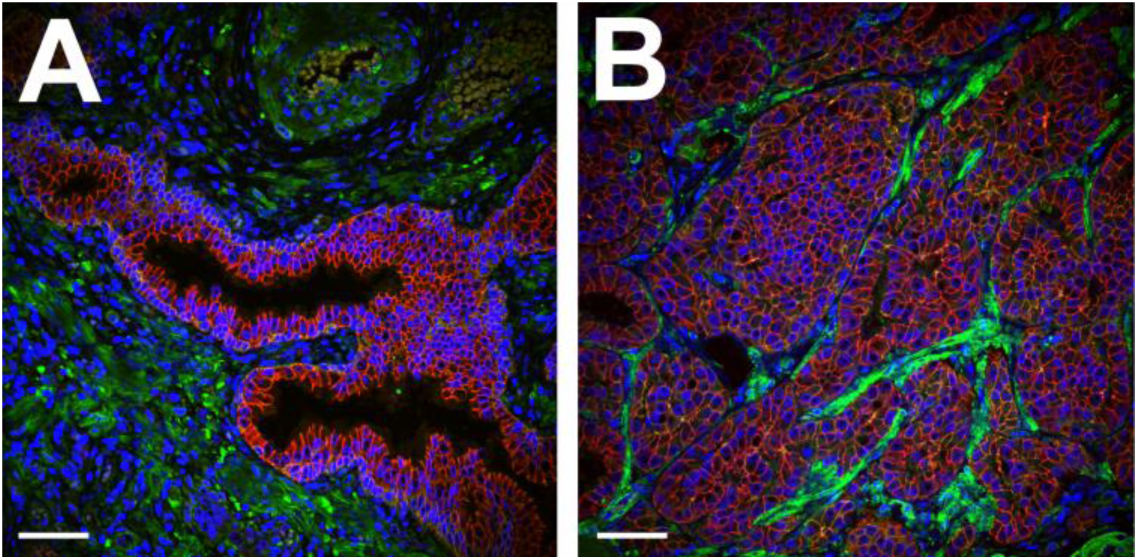
Immunofluorescence image of tissue used to derive UCSF-PR2. Figure legend: E-cadherin (cell-cell adhesion marker, red) vinculin (focal adhesion marker, green), DAPI (nuclear marker, blue). (A) Benign prostatic tissue (used for organoid UCSF-PR2). (B) High grade prostate tumor (Gleason Score 5+4), observe higher expression of E-cadherin. Scale bars equal 30 µm.

It is conceivable that PC that is different histologically would require a different concentration of Y-27632 ROCK inhibitor, which could also provide clues for therapeutic approaches. Low grade PC tumors (Gleason Score 3) convert to normal epithelium when cultured *ex vivo* as organoid cells (data not shown) or tissue (personal communication). As much as such results seem unlikely, they might indicate that early-stage PC might be a purely histological phenomenon, which reverses when the hypoxia-inducible factor 1 α (HIF1α) and the glycogen synthase kinase 3 (GSK3) are downregulated; they also suggest a strategy to reverse the transformations occurring in an aging prostate. Distinctly, GSK3α is elevated in low Gleason score tumors, while GSK3β is overexpressed in high Gleason Score tumors (Duda et al., 2020) and the isoform-specific differentiation suggests different treatment approaches. GSK3 has been demonstrated as a positive regulator in AR transactivation and PC growth independent of the Wnt/β-catenin pathway. Different types of GSK3 inhibitors including lithium show promising results in suppressing tumor growth in different animal models of PC (Li et al., 2015). High grade PC tumors (Gleason Score 4, 5) do not propagate *ex vivo* as most terminally differentiated primary tumors are dormant (Bill-Axelson et al., 2018). Terminally differentiated primary tumor cells divide once or twice per year (Thadani-Mulero et al., 2014) and are quickly overgrown by normal prostate epithelial cells, as the organoid medium is tailored for culturing of normal cells. In this context, identifying the optimal medium composition for culturing of primary PC organoids will effectively equate to providing a treatment strategy for localized disease.

#### Androgen-independent PC

Although early-stage PC can be treated reasonably well clinically, advanced forms of this disease are very aggressive and difficult to manage with existing therapies. Consequently, these tumors are associated with a high degree of morbidity and mortality. In the metastatic setting, assays to detect CTCs in the peripheral blood of cancer patients have been used clinically to provide predictive and prognostic information (Sung et al., 2013; Sung et al., 2012; Tasaki et al., 2014). Even if the use of microfluidic devices to isolate CTCs from peripheral blood for diagnostics and disease progression prognosis has gained popularity (Cristofanilli M., et al., 2004; Yu et al., 2011), such approaches carry the risk of missing CTCs during the cell capture stage. An alternative is to utilize platforms that comprise imaging all cells, without enrichment, similar to the “no cell left behind” approach (Cho and al., 2012), which had a slogan borrowed from the American administration regarding education at the time of conceiving the method. Surprisingly, the approach relied on large size CTCs only, the so-called HD (standing for “high definition”) CTCs, which were considered to be the detections of a high level of certainty, and disregarded, for example, cells with diameter similar to that of leukocytes, thus generating a sizable number of false negative selections. The reason behind this size determination was the assumption that the tumor cells present in circulation are larger than normal prostate or other epithelial cells detected in circulation and two-fold larger than leukocytes (Cho and al., 2012). While this assumption might hold true for many (but certainly not all) of the CTCs while the disease still responds to ADT, it is unlikely to be true once the tumors are castrate-resistant and the CTCs (Vid. 1) become androgen independent.

To avoid the effects of therapy and acquire resistance to androgen inhibitors, prostate tumors bypass the need for androgen (Bander et al., 2005), which affects their metabolic and cell growth patterns. While undetectable via AR expression, such CTCs can be detected by the expression of prostate-specific membrane antigen (PSMA), a cell surface marker present and enriched in 75%–95% of mCRPC (Vlachostergios et al., 2021). Such androgen-independent tumor cells have differential regulation and are much smaller in size (Fig. 5). While leukocytes are around 5.2 µm in diameter (Fig. 5A), the CTCs of a mCRPC patient can be about just 27% larger or around 6.6 µm in diameter (Fig. 5B). These results, albeit not statistically representative because of their rare event nature, indicate that in a peripheral blood draw collected from a mCRPC patient, large CTCs (of 7-13 µm in diameter or larger (Fig. S2)) could be completely absent, yet the disseminated disease might be present through a very few significantly smaller (<7 µm in diameter) tumor cells. These small CTCs can be identified via quantitative microscopy methods (Matov and Modiri, 2024) and eliminated through PSMA-targeted therapy (Unterrainer et al., 2024). In addition, mCRPC tumors often bypass the functional requirement for AR through epigenetic cellular plasticity, evidenced by the manifestation of neuroendocrine transdifferentiation (Davies et al., 2018). FGF signaling can bypass a requirement for AR, and FGF and MAPK pathways are active in metastatic AR-null PC (Bluemn et al., 2017). MAPK targeting might be efficacious in inducing ferroptosis (Wang et al., 2023) in prostate tumors with an AR-null phenotype for *TP53*-altered patients, for which the time to an endpoint of no longer clinically benefitting from AR pathway inhibitors and taxanes is comparable (De Laere et al., 2024).

**Figure 5.**
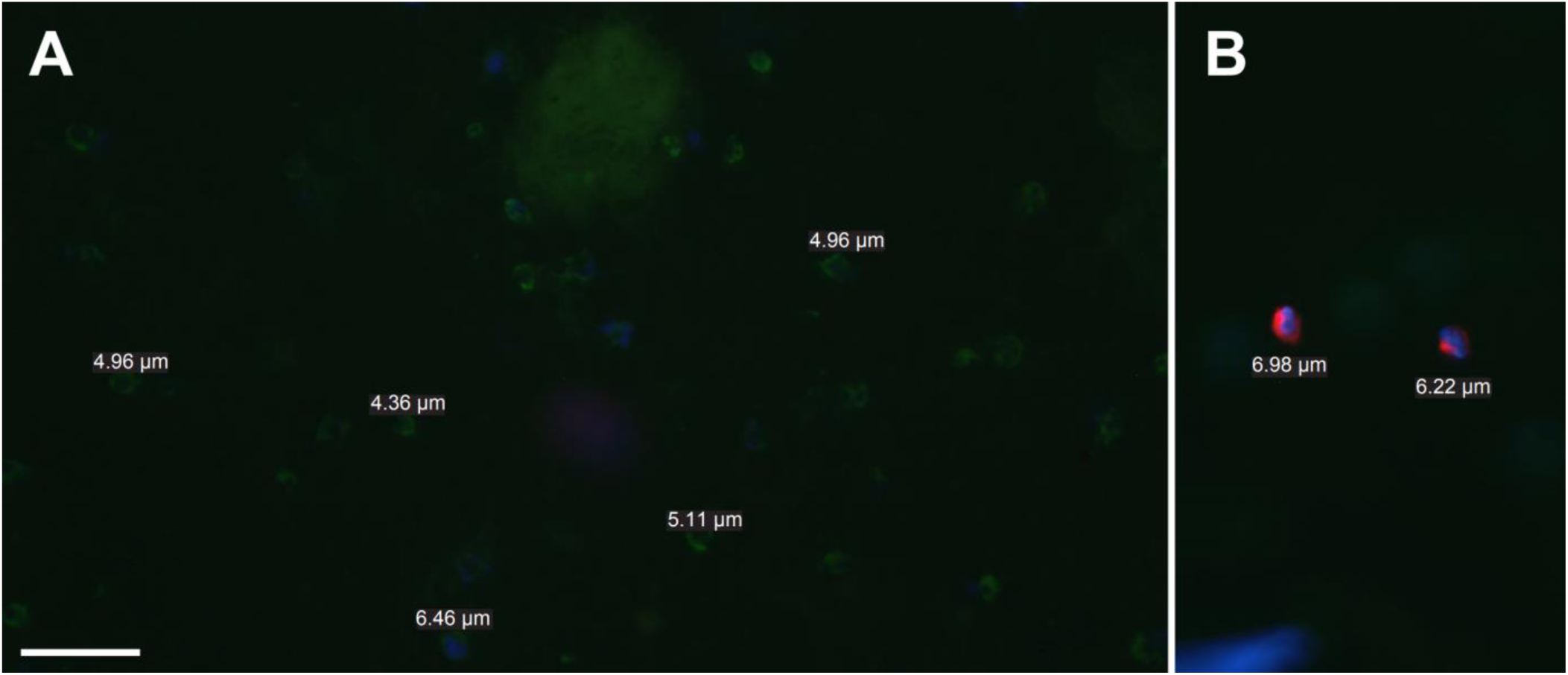
CTCs with diameter smaller than that of a normal prostate epithelial cell appear when the disease progresses to become androgen independent (or mCRPC). Figure legend: PSMA (J591, tumor marker, red), CD45 (leukocyte marker, green), DAPI (nuclear marker, blue). The scale bar equals 20 µm. (A) The diameter of patient leukocytes is between 4.3 µm and 5.2 µm. (B) The diameter of patient CTCs is as small as 6.2 µm – 7.0 µm after the disease becomes castrate-resistant.

Although not frequently mutated in PC, Ras isoforms play a pivotal role in multiple pathways that have been implicated in PC progression to androgen independence, including growth factor and cytokine induced activation of AR and its co-regulators by post-translational modifications (Whitaker and Neal, 2010). Both tubulin and actin subunits polymerize head-to-tail, thus creating polar filaments. Cytoskeletal protein complexes can be several dozen micrometers long and reach opposite sides of a cell. However, the molecular subunits of the filaments are in the nanometers range. For this reason, cellular structures can undergo rapid structural reorganizations by depolymerizing filaments at one site while polymerizing elsewhere in the cell. Actin filaments are polymers of actin monomers, and MTs consist of α/β tubulin dimers. The non-equilibrium intracellular dynamics of both filamentous actin (F-actin) and MTs, are largely determined by energy derived from ATP or GTP hydrolysis, and polymerizing MTs consist of GTP-bound tubulin dimers at their growing tips; hydrolysis follows, and the MT lattice consists of GDP-bound tubulin. Ras is activated by GTP loading and deactivated upon GTP hydrolysis to GDP. Therefore, healthy MTs would compete with oncogenic Ras for GTP, and one could indirectly limit Ras’ activity by restoring the normal functioning of the MT cytoskeleton (Basseville et al., 2015), which is dysregulated in cancer (Ertych et al., 2014; Galletti et al., 2014; Gonçalves et al., 2001; Harkcom et al., 2014; Thoma et al., 2010) and regulated by functional pVHL (Thoma et al., 2010). Ras-driven cancers have proven very difficult to treat (Hong et al., 2020; Stephen et al., 2014; Zhang et al., 2018), and the most common *KRAS* p.G12C mutation can be targeted via proteins fused to the pVHL E3 ubiquitin ligase (Bery et al., 2020; Bond et al., 2020). Using proteolysis targeting chimeras to degrade proteins linked to tumorigenesis has emerged as a potential therapeutic strategy for cancer (Khan et al., 2020). These heterobifunctional molecules consist of one ligand for binding to a protein of interest and another to an E3 ligase, connected via a linker and only a few E3 ligases can be used to degrade target proteins with this approach, including pVHL (Zhao et al., 2019). In PC, proteolysis targeting chimera agents that utilize pVHL E3 ligase ligands (Han et al., 2019) represent promising AR-targeting therapeutics and could become an important part of disease treatment (Zang et al., 2024) as oral protein degradation agents have shown encouraging results in clinical trials (Qi et al., 2021).

Op18, a MT polymerization inhibitor (Cassimeris, 2002), has been associated with taxane resistance in androgen-independent PC (Mistry and Atweh, 2006). In meiotic spindles, we discovered that upregulation of Op18 leads to the formation of about three-fold smaller spindles (Houghtaling et al., 2009) and it inhibited the formation of a particular subset of MT bundles that directly link the two spindle poles (termed non-kinetochore MTs). The non-kinetochore MTs represent a distinct MT subpopulation and polymerize at slightly higher rates (about 0.3 µm/min faster) than those MT bundles that attach to the kinetochores, which have their dynamics tightly regulated by the protein complexes at the kinetochore (Matov, 2025b; Vallotton et al., 2003). We measured a dose-dependent decrease in spindle size upon addition of up to 2.5-fold excess Op18 (endogenous Op18 concentration of 6 μM (Belmont and Mitchison, 1996), 15 μM of recombinant Op18 added) without significantly reducing MT polymerization rates (<1 µm/min attenuation) (Houghtaling et al., 2009; Matov, 2025a). In interphase cells, our analysis of gene expression data of four PC patient organoids refractory to androgen therapy (Fig. S3) indicated upregulation of Op18 in the patient organoid for which we measured lower MT polymerization rates (>3 µm/min attenuation) (Matov, 2024d). Taken together, the data indicates that in androgen-independent PC tumors the upregulation of Op18 might induce a reduction of cell size and resilience to therapeutic interventions. Overall, our work has identified differential mechanisms of cytoskeletal remodeling in epithelial tumors leading to either very large spindles or very small spindles in cells that are drug resistant.

#### Small molecules’ effects on MT dynamics

An important process for tumor development and metastasis involves the remodeling of the 3D microenvironment surrounding the tumor cells. Dynamic remodeling of the tumor microenvironment is a critical, prerequisite step for metastatic tumor growth driven by signaling between cancer cells and cells in the surrounding extracellular matrix. MT and cytoskeleton reorganization are factors regulating the signaling processes that promote the remodeling of the tumor microenvironment and drug resistance. For example, the fusion of TMPRSS2 with ETS genes is a recurrent genomic alteration in PC and the ETS-related gene (ERG) is the most common fusion partner for the androgen regulated gene TMPRSS2 (Tomlins et al., 2005). ERG affects several parameters of MT dynamics and inhibits effective drug-target engagement of docetaxel or cabazitaxel with tubulin (Matov et al., 2013). In CTCs captured from an ERG-negative patient, we observed robust drug-target engagement in 41 of the 66 captured CTCs (62%) and in 82 of the 105 captured CTCs (78%) following docetaxel and cabazitaxel treatment, respectively (Galletti et al., 2014). In contrast, in the CTCs isolated from an ERG-positive patient, there was no increase in MT bundling with either treatment and MT bundle formation is the result of drug-mediated MT polymer stabilization, and as such it represents a hallmark of taxane cellular activity (Tasaki et al., 2014). Next, to evaluate the effects of metastatic, receptor-negative epithelial tumors on the MT cytoskeleton, we measured MT polymerization dynamics using automated tracking of EB1/3ΔC-2xEGFP comets (Harkcom et al., 2014; Kumar et al., 2009; Matov et al., 2016; Matov et al., 2013; Matov et al., 2005; Thoma et al., 2010). We have shown that there exists a rocking pendulum-like motion in many of the mitotic spindles in MDA-MB-231 cells (Matov, 2024d). The MDA-MB-231 cells have the largest spindles and, while most of the spindles in other cell lines remain stationary until anaphase, many of them rotate at up to an angle of ±40° (Matov et al., 2015) to the left and right of the polar spindle axis. This rapid rocking motion over an 80° angle is reflected in the increased EB1 velocity. Altered MT dynamics endow cancer cells with both survival and migratory advantages (Mitchison, 2012), which can be correlated with drug resistance (Gonçalves et al., 2001). In interphase cells, our results show that a 2-hour treatment with 100 nM paclitaxel brings MT dynamics to a very uniform behavior (Tran et al., 2013; Tran et al., 2012) by suppressing the fast polymerizing MTs and lowers significantly the overall polymerization rates (Fig. 6). Tracking of EB1 comets in a MCF10A normal breast epithelium cell line indicated that the EB1 speeds in cells from normal breast epithelium move about 30% to 70% faster in comparison to breast cancer cells (see Fig. S4 and (Matov, 2024d)).

**Figure 6.**
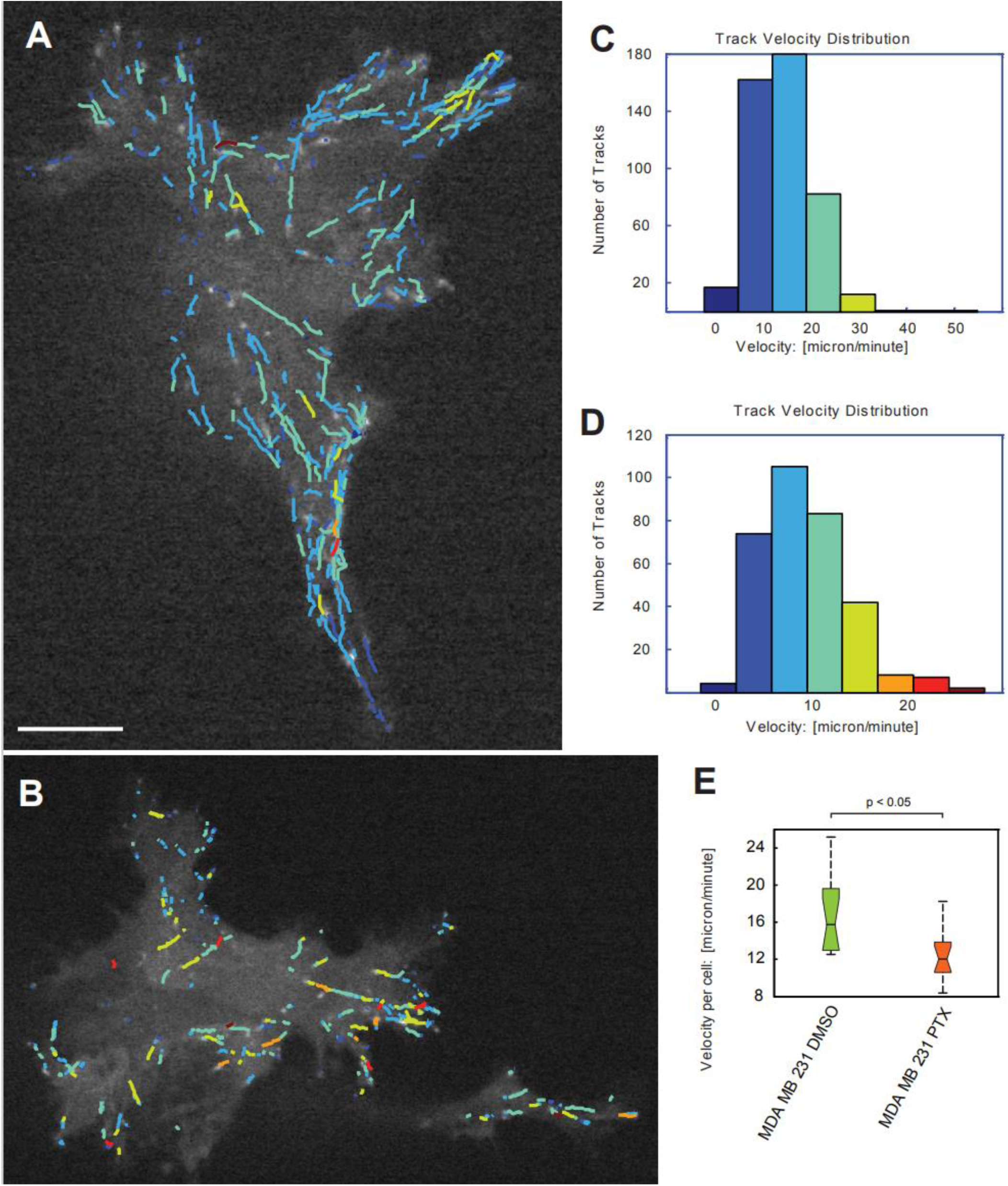
Paclitaxel attenuates MT growth rate in breast cancer (MDA-MB-231) cells. MDA-MB-231 cells were transiently transfected with EB1ΔC-2xEGFP and treated with vehicle (DMSO) or 100 nM paclitaxel for 2 hours. EB1ΔC-2xEGFP comet time-lapse sequences were recorded for every 0.5 seconds for 125 frames and computationally tracked (Yang et al., 2005). The color-coding represents EB1 speeds and colder colors correspond to lower speeds, and warmer colors correspond to faster speeds. The scale bar equals 10 µm. Overlay of EB1 comets raw data and computer-generated growth tracks with a minimum lifetime of four frames (or two seconds) is shown on (A) for DMSO, for which the median speeds were about 11 µm/min and the maximal speeds were about 49 µm/min and (B) for paclitaxel, for which the median speeds were about 8 µm/min and the maximal speeds were about 25 µm/min. Histograms of growth velocities in µm/min are shown on (C) for DMSO and (D) for paclitaxel. (E) Distributions of growth speed of 155,981 EB1 comets in 14 DMSO treated (left) and 39,989 EB1 comets in 14 paclitaxel (PTX) treated cells (right) and of per cell mean velocity. Significance of the difference between concentrations was determined by a permutation *t*-test (p < 0.05).

Besides differential regulation in disease, the size of the cells and the mitotic spindles they form also affects MT dynamics. We have shown that higher EB1 density decreases MT growth rates (Geisterfer et al., 2020; Jevtic et al., 2014; Jevtic et al., 2013), which can lead to taxane resistance as well (Galletti et al., 2014; Matov, 2024d). Taken together, these data suggest that smaller tumor cells are also more likely to be resistant to therapy.

Nicotinamide adenine dinucleotide (NAD+) is an endogenous small molecule that has effects on diverse processes, including obesity, lifespan, and cancer, which can increase MT polymerization rates (Harkcom et al., 2014) and restore normal MT regulation and homeostasis, which suggests its utilization during the treatment of tumors of epithelial origin. However, there have thus far been no studies on the effects of the essential redox metabolite NAD+ specifically in PC. NADH elevation during chronic hypoxia leads to pVHL-mediated HIF1α degradation (Joo et al., 2023), which has metabolic consequences for quiescent cells. HIF1α drives resistance to ferroptosis in solid tumors (Yang et al., 2023) and a transient increase of HIF1α during the G1 phase prevents the onset of starvation-induced apoptosis (Belapurkar et al., 2023). GSK3β also plays a role in modulating ferroptotic sensitivity by regulating cellular iron metabolism (Wang et al., 2021). In bacterial antibiotic resistance, a non-mutational mechanism of acquired drug resistance is the state of persistence (Fisher et al., 2017) during which a small population (<5%) of the cells become quiescent. In cancer, the quiescent surviving persister cells minimize the rates of proliferation and, thus, become not susceptible to treatment with mitotic inhibitors and targeted therapies (He et al., 2024; Russo et al., 2024). This reversible process of becoming drug-tolerant (Green, 2024) is linked to upregulation of mesenchymal markers and downregulation of epithelial markers in the persister cells. During drug holiday, the cells revert to normal metabolism and expand their population, while becoming again susceptible to drug treatment. This cycle repeats multiple times during patient treatment. While the tumor is under drug pressure, however, the few drug-tolerant persister cells pay a metabolic price and become temporarily vulnerable to GPX4 inhibition (Hangauer et al., 2017). We have investigated the effects of small molecules, RSL3 and ML210, which inhibit the activity of GPX4 and induce ferroptosis in, initially drug-sensitive, patient cells in PC organoids with non-mutational acquired drug resistance (Matov, 2024a) and identified a clinical approach to impede disease relapse and eliminate residual disease (Hangauer et al., 2017). Ferroptosis has a dual role in cancer. It plays a role in tumor initiation, tumorigenesis, which depends on inflammation-associated immunosuppression triggered by ferroptotic damage (Chen et al., 2021) and later, during treatment, in tumor suppression (Hangauer et al., 2017). Recently, we identified the presence of patterns of DNA fragmentation linked to a decrease in the number of caspase-dependent DNA cleavage fragments with lengths 198 bp, 364 bp, and 521 bp, and differential detoxification of ROS and metabolic/redox patterns in colorectal cancer (Matov, 2024b). NAD+ promotes cellular oxidative metabolic reactions and enables mitochondrial respiration to support cell growth and survival (Griffiths et al., 2020). Taken together, these results suggest the utilization of NAD+ and other endogenous molecules in the treatment of PC in the context of modulating the drug susceptibility of resistant PC, i.e., as drug regimen sensitizing compounds.

#### GSK3β and pVHL maintain MT dynamicity

Mutations in the *VHL* gene lead to various patterns of MT dynamics signatures (Thoma et al., 2010). pVHL regulates the activity of the HIF1α (Genbacev et al., 2001; Kaelin and Ratcliffe, 2008; Maxwell et al., 2001; Staller et al., 2003), and *VHL* mutations lead to a variety of malignant and benign tumors of the eye, brain, spinal cord, kidney, pancreas, and adrenal glands. Hypoxic microenvironment drives the selection of aggressive cancer cells through actin cytoskeleton polarization, downregulation of E-cadherin, and epithelial-mesenchymal transition (Buart et al., 2021). Mutations in *VHL* lead to an increased GTP hydrolysis (Thoma et al., 2010) and dysregulated MT dynamics (Barry and Krek, 2004), a decreased drug susceptibility, and increased oncogenic activity. Dysfunctional pVHL tumor suppressor leads, for instance, to upregulation of the MT-associated protein MAP1, which has been implicated in pathogenesis (Czaniecki et al., 2019; Fifre et al., 2006) as well as drug resistance in several cancers (Laks et al., 2018) and, thus, represents a target within the MT transcriptome to sensitize resistant disease. We have demonstrated that it is possible to associate dysregulation of specific aspects of MT dynamics to mutations occurring within a particular domain of pVHL (Thoma et al., 2010). Some tubulin inhibitors bind to individual tubulin dimers and block tubulin polymerization (MT-destabilizing drugs); others bind on the luminal side of the MT lattice and inhibit MT‘s ability to depolymerize (MT-stabilizing drugs). Mutations in the MT-interacting/HIF1α-interacting domain are more often linked to changes in MT polymerization/depolymerization (or turnover) rates (e.g., Y98H mutation), and therefore, such disease could be treated with MT-destabilizing drugs. *VHL* mutations in the Elongin C-interacting domain are more often linked to changes in the MT catastrophe and rescue frequencies (e.g., R161P mutation), and therefore, such disease could be treated with MT-stabilizing drugs.

Signaling through HIF1α is dependent on interphase MTs and the protein accumulation and translational regulation of HIF1α is inhibited by taxane treatment (Carbonaro et al., 2012; Carbonaro et al., 2011). Activation of quiescent fibroblasts by tumor cells is dependent on the integrity of tumor cell MT cytoskeleton and, thus, taxane treatment reduces fibroblast activation as a result of drug-mediated suppression of MT dynamics (Tran et al., 2013; Tran et al., 2012). We also identified an isoform-specific attenuation of MT polymerization rates in *VHL*^-/-^ renal cell carcinoma cells in which either the short or long pVHL isoforms were reconstituted. Viral reconstitution of pVHL19 partially reduced the EB3 comet speeds (with 2.1 µm/min) and increased the times of MT pause with one second on average, while pVHL30 further reduced the EB3 speeds (with 4.1 µm/min) but increased the times of MT pause with just half a second on average (Thoma et al., 2010). Therefore, the overall attenuation of MT polymerization rates for both pVHL isoforms was very similar, but demonstrated differential isoform-specific mechanisms, which may represent a personalized targeting approach. In addition, we identified the mechanism by which pVHL30 protects MTs from depolymerization after treatment with 40 nM nocodazole; while MT tips in *VHL*^-/-^ renal cell carcinoma cells instantly get sparser upon adding the drug (Vid. 2) and within six minutes depolymerized when co-cultured with the drug, in cells with reconstituted pVHL30, MTs paused significantly longer and this way resisted the drug treatment.

Further, analysis of pVHL species carrying nonphosphorylatable or phosphomimicking mutations at Ser-68 and/or Ser-72 revealed a central role for phosphorylation events in the regulation of pVHL’s MT stabilization (Hergovich et al., 2006). Our measurements of MT dynamics showed that in cells stably transfected with retroviral vectors expressing the pVHL-GSK3β phosphorylation-site mutants (S68/72A and S68/72D) different alteration in MT homeostasis have been induced (Fig. 7). pVHL mutant S68/S72A (linked to cilium rescue when GSK3β is subjected to inhibitory phosphorylation (Thoma et al., 2007a)), which is resistant to phosphorylation by GSK3β, leads to reduced MT growth dynamics of 15.8 µm/min (Table 1) and increased MT pausing after 40 nM nocodazole treatment, similar to the effects of reconstituting the long pVHL isoform. GSK3β and pVHL cooperatively maintain primary cilia (Davenport and Yoder, 2005) in RCC-4 cells; pVHL plays a key role in maintaining the structural integrity of the primary cilium, a MT-based cellular antenna important for suppression of uncontrolled proliferation of kidney epithelial cells and cyst formation. The cilium maintenance function of pVHL is independent of its role in HIF1α degradation, but it is dependent on its ability to stabilize the MT network of the ciliary axoneme (Thoma et al., 2007a). pVHL-negative cells lose their primary cilia in response to serum stimulation. The effect of serum on pVHL-negative cells is dependent on PI3K signaling and could be mimicked by inhibition of GSK3β. Thus, pVHL and GSK3β demonstrate a genetic redundancy with respect to cilia maintenance; inhibition or loss of either protein alone has no effect on primary cilium, but inhibition or loss of both causes cells to rapidly lose their cilia (Thoma et al., 2007b). Overall, our analysis shows that growth factor deprivation enhances MT turnover and destabilizes MT, and that GSK3β-mediated phosphorylation and inhibition of pVHL function induces these changes in MT homeostasis.

**Figure 7.**
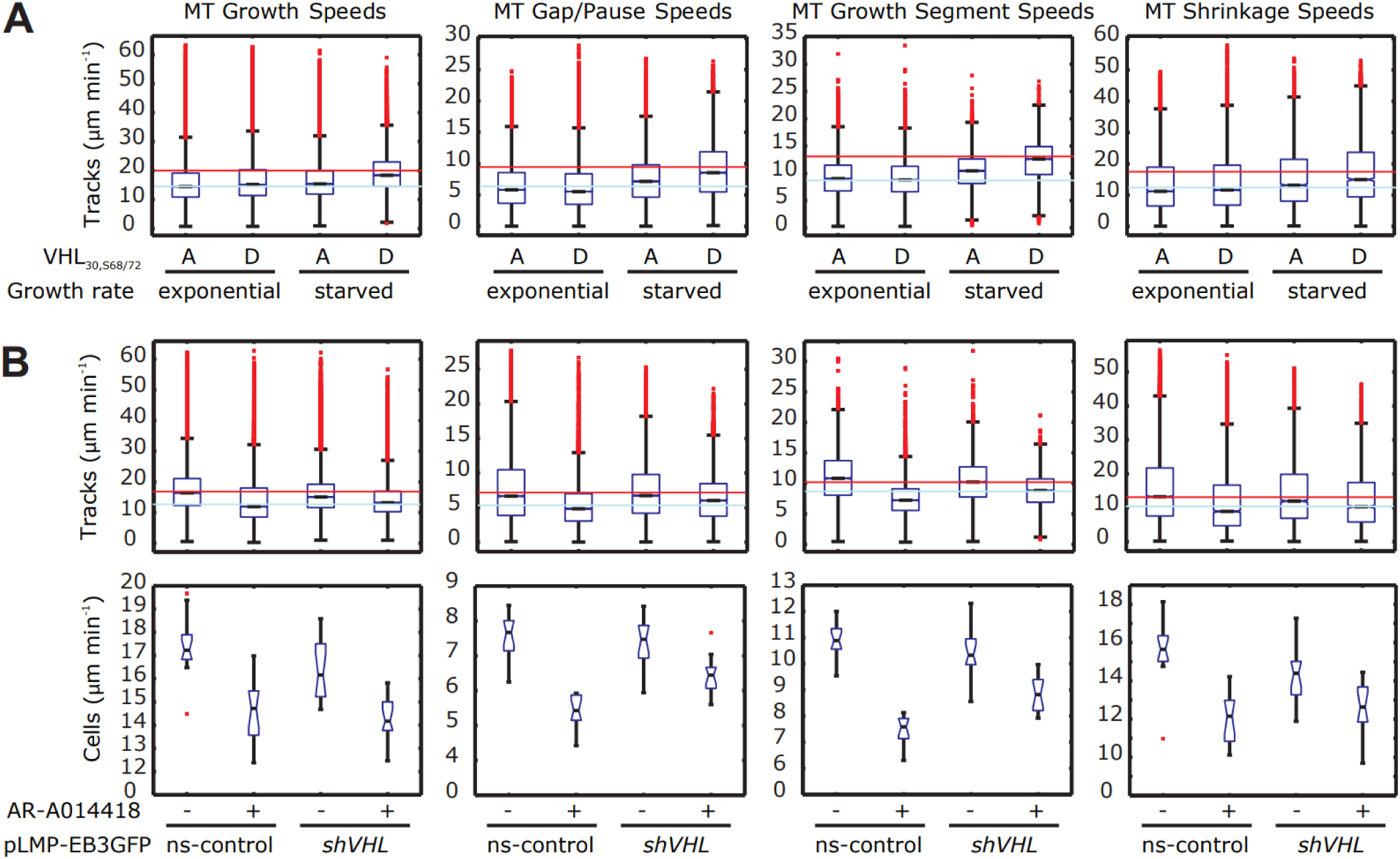
pVHL and GSK3 affect MT dynamics in interphase and quiescent cells. MT tips are labeled with EB3ΔC-2xEGFP, imaged for a minute (with an acquisition rate of two images per second) and computationally analyzed (Matov et al., 2010). About a dozen cells were analyzed per condition containing about 4,000 MT tracks each with a minimum track lifetime of 2 seconds. The four MT dynamics parameters (Matov et al., 2010) compared are as follows: (1) MT polymerization rates (growth speeds), (2) MT dynamics during trajectory gaps, when the EB3ΔC-2xEGFP molecular marker is not visible (pause/gap speeds), (3) MT polymerization rates over multiple trajectories interrupted by periods when the EB3ΔC-2xEGFP molecular marker is not visible (growth segment speeds), (4) MT depolymerization rates (shrink/shortening speeds). (A) Box plots representing the measured MT-dynamic parameters in RCC-4 cells reconstituted with the phosphorylation site mutant pVHL30(S68/72A) resistant to phosphorylation by GSK3β and the phosphorylation-mimicking mutant pVHL30(S68/72D) under exponential growth and serum-starved conditions. For comparison, each parameter’s median value is indicated for RCC-4 cells reconstituted with pVHL30 under exponential growth (light blue line) and serum-starved (thin red line) conditions. (B) Box plots representing the measured MT-dynamic parameters per track (upper panels) and per cell (lower panels) of a non-silencing control (ns-control) RPE-1 cells and cells depleted of pVHL using retroviral expression of microRNA based small hairpin RNA (shRNAmir) (via cloning into pLMP-EB3GFP retrovirus vector (Hergovich et al., 2006)) against *VHL* (*shVHL*) under serum-starved conditions. To get the cells in a quiescent state they were grown for 24 hours and then serum-starved for 24 hours. For GSK3 inhibition, 20 hours of starvation was followed by incubation with 100 μM AR-A014418 for 4 hours in the same medium. For comparison, each parameter’s median value for MT tracks is indicated for a non-silencing control (ns-control) RPE-1 cells (light blue line) and cells depleted of pVHL using retroviral expression of microRNA based small hairpin RNA under exponential growth conditions (thin red line). Per cell measurements are added because the cell-to-cell variation is the single biggest source of variation in the median value of a MT parameter. The decrease in the values for each parameter after the addition of AR-A014418 was statistically significant.

**Table 1.** Loss of pVHL leads to MT turnover rates (in red) similar to quiescent cells with phosphorylated pVHL. Oppositely, in cells with reconstituted pVHL, MTs dynamics (in blue) are similar to quiescent cells with inhibited GSK3. Computational analysis of MT dynamics performed using ClusterTrack (Matov et al., 2010). The cell lines consist of clear cell renal cell carcinoma (ccRCC) cells and hTERT-immortalized retinal pigment epithelial (RPE-1) cells (Vid. 3). RCC-4 cells, which are deficient for pVHL, were engineered to stably produce the empty vector as control (pCMV(R)-VHL^-/-^), the long or short pVHL isoforms (pCMV(R)-VHL30 and pCMV(R)-VHL19, respectively) and the phosphorylation site mutant pVHL30(S68/72A) that is resistant to phosphorylation by GSK3β or the phosphorylation-mimicking mutant pVHL30(S68/72D). RPE-1 cells, which express wild-type pVHL, were transfected with retroviral vectors expressing a non-silencing (ns-control) control or a miRNA30-based short-hairpin RNA to knock-down pVHL (shVHL) (Vid. 4). To get in a quiescent state, cells were serum-starved for 24 hours. The GSK3 inhibitor used was VIII (AR-A014418) at concentration of 100 μM. For the 12 conditions, a combined number of 173 movies containing over 640,000 MT tracks with duration of at least 2 seconds were analyzed. Each movie consists of 125 image frames taken 0.5 seconds apart. Cilia maintenance data is incomplete for five of the conditions (serum-starved cells). The 10^th^ condition (RCC-4 serum-starved cells expressing the long isoform of pVHL) is based on only six videos (with one or two cells) and fewer MT tracks than normally detected per video (datasets of at least either 10 videos or 20,000 MTs are considered), and some of the cells were either much smaller than in any other condition or represented an unusual morphological outlier; these factors are indications of a statistically not representative dataset and poor data quality and, thus, the analysis result constitutes an artifact.

| Cells | Mutation | Type | MT tip speed | Videos | MT tracks | Cilia |
| --- | --- | --- | --- | --- | --- | --- |
| RCC-4 | null | Loss of pVHL | 18.7 $\mu\text{m}/\text{min}$ | 10 | 37,120 | No |
| RCC-4 | none | Long isoform | 14.8 $\mu\text{m}/\text{min}$ | 9 | 41,844 | Yes |
| RCC-4 | none | Short isoform | 16.8 $\mu\text{m}/\text{min}$ | 12 | 28,578 | Yes |
| RCC-4 | S68/72A | GSK3 non-phos. | 15.8 $\mu\text{m}/\text{min}$ | 30 | 166,669 | Yes |
| RCC-4 | S68/72D | GSK3 phosphor. | 16.7 $\mu\text{m}/\text{min}$ | 28 | 102,202 | No |
| RPE-1 | ns | Long isoform | 15.4 $\mu\text{m}/\text{min}$ | 12 | 68,939 | Yes |
| RPE-1 | shVHL | Loss of pVHL | 17.6 $\mu\text{m}/\text{min}$ | 13 | 55,286 | No |
| RPE-1 | ns<br>s-starved | Long isoform<br>100 $\mu\text{M}$ DMSO | 17.6 $\mu\text{m}/\text{min}$ | 15 | 39,541 | n.a. |
| RPE-1 | ns<br>s-starved | 100 $\mu\text{M}$ GSK3<br>inhibitor VIII | 14.6 $\mu\text{m}/\text{min}$ | 13 | 35,515 | n.a. |
| RCC-4 | s-starved | Long isoform | 20.3 $\mu\text{m}/\text{min}$ | 6 | 8,590 | n.a. |
| RCC-4 | S68/72A<br>s-starved | GSK3 non-phos. | 16.7 $\mu\text{m}/\text{min}$ | 13 | 34,778 | n.a. |
| RCC-4 | S68/72D<br>s-starved | GSK3 phosphor. | 19.3 $\mu\text{m}/\text{min}$ | 12 | 22,222 | n.a. |

#### MT regulation in quiescent cancer cells

Phosphorylation of pVHL at Ser-68 proceeds mainly in serum-starved cells and is consistent with the fact that GSK3β is predominantly active under conditions of serum starvation (Lochhead et al., 2006). Our measurements of EB3 comet speeds after perturbations of the GSK3β−pVHL axis indicated altered MT dynamics instability. Specifically, serum-starvation of pVHL-positive RPE-1 cells altered MT turnover to the same extent as shRNA-mediated depletion of pVHL in exponentially-growing cells when serum was present (Fig. 7, Table 1). Importantly, inhibition of GSK3β in serum-deprived cells attenuated MT turnover, which reached rates similar to those we measured in exponentially-growing, pVHL-positive RPE-1 cells. To achieve GSK3 inhibition, 20-hour starvation was followed by a 4-hour incubation with 100 μM GSK3 inhibitor VIII (AR-A014418), which is used in the treatment of PC (Li et al., 2015). Our results suggest that the state of quiescence is characterized by increased MT dynamics, as compared to the state of exponential growth, and that GSK3β plays a critical role in mediating the changes in MT dynamics (Table 1). To assess whether GSK3β-mediated phosphorylation of pVHL contributes to the measured changes in MT dynamics associated with quiescence, we analyzed serum-starved RCC-4 cells producing either the phosphorylation site mutant pVHL30(S68/72A) that is resistant to phosphorylation by GSK3β or the phosphorylation-mimicking mutant pVHL30(S68/72D) (Fig. 7, Table 1). Serum deprivation caused an increase in MT turnover in RCC-4 cells that expressed pVHL30 or pVHL30(S68/72D), but not in cells expressing pVHL30(S68/72A). Changes in MT dynamics such as an increase in MT turnover upon serum-deprivation were coupled with a decrease in MT shrinkage times, i.e., an increase in MT catastrophe frequencies. Our results suggest that phosphorylation of pVHL at Ser-68 by GSK3β inactivates the MT stabilization functions of pVHL in terms of attenuation of MT turnover and inhibition of catastrophe events and indicate that phosphorylation is a critical aspect of the regulation of MT dynamics when cells enter a quiescent state.

#### MTs in metastatic cells

The cytoplasmic linker protein CLIP170, by recognizing conformational changes, like EB1/3, binds growing MT tips and forms comets (Vid. 5) and has been linked to taxane resistance (Thakkar et al., 2021). In migrating epithelial cells, such as cells undergoing epithelial-mesenchymal transition, cytoplasmic linker-associated proteins (CLASPs) bind (or track) selectively MT growing tips in the cell body but bind along the MT lattice in the lamella. GSK3β phosphorylates cytoplasmic CLASPs at multiple sites in the domain required for MT growing tip interaction. Expression of constitutively active GSK3β destabilizes lamella MTs by disrupting lateral MT interactions with the cell cortex.

GSK3β-induced lamella MT destabilization (Kumar et al., 2009) can be partially rescued by expression of CLASP2 with mutated phosphorylation sites indicating that CLASP-mediated stabilization of peripheral MTs, which likely occurs in the vicinity of focal adhesions (FAs), is regulated by local GSK3β inactivation. Similar to the effects of functional pVHL (Thoma et al., 2010), ectopic activation of GSK3β at the cell edge results in inhibition of MT catastrophe events (Kumar et al., 2009). When MTs switch to a state of depolymerization less often, the cells become larger, the MTs get longer and remain as polymers for an extended period of time – and that leads to MT acetylation (Janke and Montagnac, 2017), which also (further) inhibits MT catastrophe events (Matov et al., 2010). MT post-translational modifications in α-tubulin that induce an increase in polymer rigidity and mechanical resistance, such as detorysination and acetylation, stabilize MTs (Eshun-Wilson et al., 2019) and also induce drug resistance (Galmarini et al., 2003; Kavallaris, 2010; Matov et al., 2010). Overall, our investigation indicates that both faster than normal MT turnover rates and slower than normal MT turnover rates are hallmarks of drug resistance.

### ANTICIPATING DRUG RESISTANCE

#### Resistance evaluation via circular Hough transform

In drug sensitive interphase cells, MT bundle formation is the result of drug-mediated MT polymer stabilization and as such it represents a hallmark of taxane cellular activity (Sung and Giannakakou, 2014). MT bundles eventually form circular shapes as cells approach apoptosis. We applied circular Hough transform to detect MT bundling and reorganization in drug-treated cells (Fig. 8). Hough transform is a computational technique, which allows the automated detection of a variety of shapes in images (Hough, 1959), most commonly used in the area of computer vision for line detection (Mattavelli et al., 2001). Such algorithms have been in use, for instance, by the Danish postal services since the onset of the century to sort mail by automatedly detecting the envelope stamp and have drastically improved the sorting efficiency and decreased the utilization of human, manual labor. Several hours after treatment with paclitaxel, A549 cells began changing their shape into a circle and formed thick tubulin rings in 83% of the cells (Fig. 8A), which indicates sensitivity to the drug action. Knockdown of the BRCA1 gene in A549 cells (Sung and Giannakakou, 2014) affects their susceptibility to treatment with paclitaxel and they become resistant (Fig. 8B). There is a variability of the cellular response in terms of morphological changes and only 27% changed their shape into a circle and formed thick tubulin rings (Fig. 8B). Further, when we compared the pixel intensity over these rings of bundled MTs in the affected cells from the parental A549 cells and the derivative BRCA1 knockdown cells, the overall brightness was 70% higher in the paclitaxel-susceptible parental cells (Fig. 9). These results indicate that even if there are measurable drug-induced changes in about a quarter (27%) of the resistant derivative BRCA1 knockdown cells, the effects of paclitaxel are significantly reduced. Overall, these data demonstrate two different mechanisms of taxane resistance.

**Figure 8.**
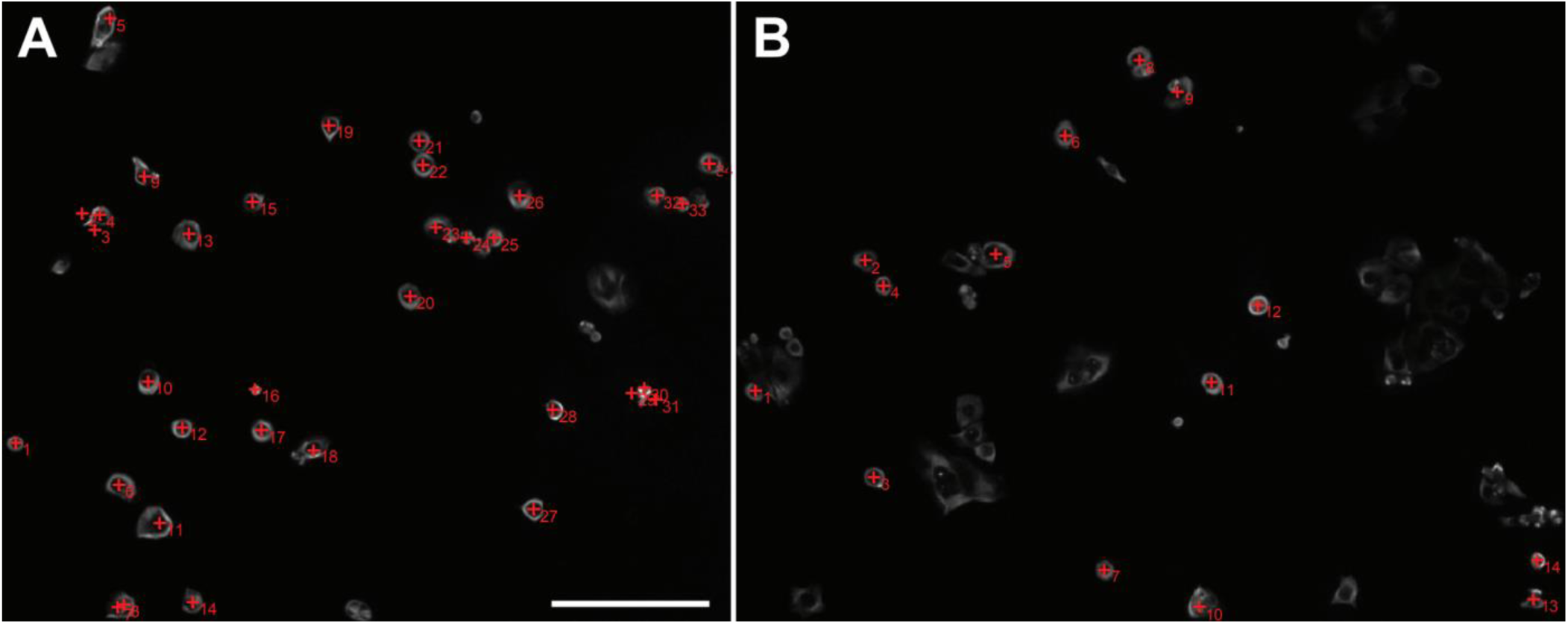
Paclitaxel resistance resulting from knockdown of BRCA1 in adenocarcinoma (A549) cells. Cells were treated with 100 nM paclitaxel for 18 hours. Cell center in drug-affected cells is marked with a red +. Cells with intensity rings in α-tubulin immunofluorescence images (Sung and Giannakakou, 2014) are detected and enumerated. Scale bar equals 100 µm. (A) A549 cells respond to paclitaxel treatment. In this field of view, 34 cells responded (83%) to the treatment and their MT cytoskeleton formed round bundles, four cells were not affected (9.7%) by the drug and did not form bundles, and three cells were already apoptotic (7.3%) by the time of imaging. (B) BRCA1 knockdown in A549 cells makes the cells resistant to paclitaxel. In this field of view, 14 cells responded (27%) to the treatment and their MT cytoskeleton formed round bundles, 30 cells were not affected (58%) by the drug and did not form bundles at all, four cells were partially affected (7.5%) but did not form clear bundles, and four cells were already apoptotic (7.5%) by the time of imaging.

**Figure 9.**
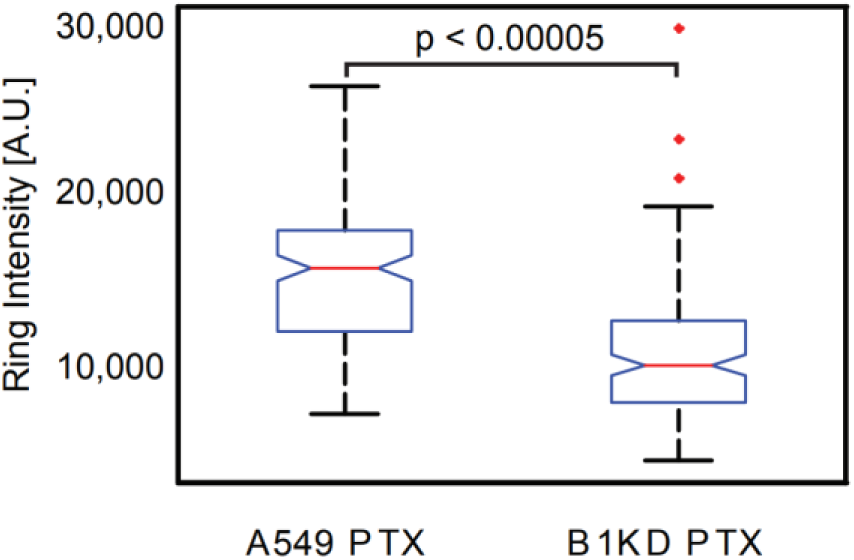
Quantification of paclitaxel (PTX) resistance resulting from knockdown of BRCA1 (B1KD) in adenocarcinoma (A549) cells. Cells were treated with 100 nM paclitaxel for 18 hours. Cells which were sensitive to the drug, formed circular tubulin bundles after treatment (see Fig. 8A for the 34 drug (83%) sensitive cells in the parental A549 cell line and Fig. 8B for the 14 (27%) sensitive cells in the B1KD cell line). Comparison of the overall pixel intensity of these tubulin rings formed after paclitaxel treatment indicates much lower drug response with less bundles formed by stabilized MTs in the B1KD cell line, for which the overall ring intensity was lower with about 7,000 (17,000 vs. 10,000) as a median value. The differential drug-induced effects in the sensitive cells of the two cell lines suggests an additional mechanism of taxane resistance and a partial drug response in the B1KD cells.

#### Correlation of live-cell MT dynamics and gene expression signatures

To validate any *ex vivo* results regarding drug response and resistance, one could correlate the MT dynamics analysis in patient-derived tumor organoids with gene expression data and clinical response of the same patients. Our objective has been to identify candidate targets amongst the proteins which regulate particular parameters of MT dynamics (Lyle et al., 2009a; Lyle et al., 2009b). To achieve this end, we will use pattern analysis algorithms (He et al., 2013; Tiemann et al., 2013) and expect to be able to match the patient gene expression alterations (Fig. S3) to the pattern of deregulation of MT dynamics in a particular tumor (Matov, 2024d). Having measured MT deregulation patterns across multiple patients, we will create profiles of susceptibility to tubulin inhibitors within a statistical framework that classifies MT trajectories by mapping gene expression patterns to patient treatment outcomes (similar to (Monnier et al., 2015)).

#### Validation of epigenetic signatures and evaluation of therapeutic interactions

Next, as MT-regulated genes implicated in drug resistance for the concrete patient are identified, organoids from knockdown experiments of inhibiting multiple of these MT regulators could be injected into mice and tested for an improved drug response (Fig. 10). We will test knockdown organoids in terms of MT dynamics and dose-response morphology and image texture metrics (Matov, 2024a) as well as for alteration in drug susceptibility in patient derived xenografts (PDX) models in this context. We tested culturing of prostate tumor tissue slices for up to 48 hours (Keshari et al., 2013) without tissue dissociation using a bioreactor (Maund et al., 2014) with the objective of developing another assay for the evaluation of drug action by computing and comparing the changes in the power spectrum of tissues after drug treatment imaged as a 3D hologram (Jeong et al., 2010). Another validation approach is to compare organoid gene expression signatures via high-throughput transcriptomics, which can evaluate the pharmacogenomics and establish biological pathway altering concentrations (Harrill et al., 2021). By utilizing multiple validation methods, we will be able to establish not only the ability of our approach to anticipate and overcome drug resistance but also to evaluate whether the use of putative compounds will induce accumulative side effects beyond the desired metabolic changes for drug efficacy (Phillips et al., 2018).

**Figure 10.**
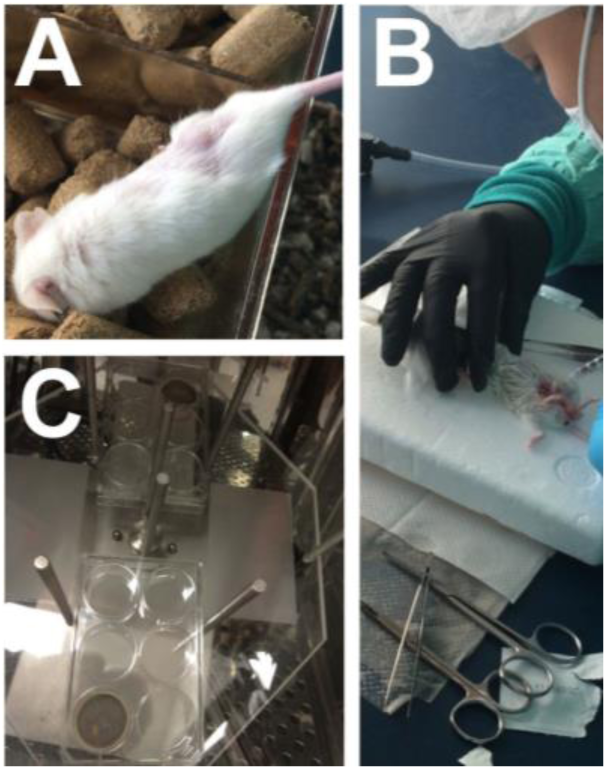
Injecting PC organoids to establish patient derived xenografts in PC and culturing PC tissue *ex vivo*. (A) Tumor tissue from a radical prostatectomy case was planted subcutaneously by a surgeon in an immunodeficient mouse. (B) Injecting metastatic PC organoids into an immunodeficient mouse. (C) Culturing PC tumor tissue on a (bioreactor) shaker. Upper right and lower left wells contain slices of tissue from 1 mg punch core biopsy with Gleason Score 5+4 and 80% tumor density to evaluate drug action by computing the changes in the power spectrum of tissue of a 3D hologram (Jeong et al., 2010) after treatment with tubulin inhibitors.

## CONCLUSIONS

About a third of the patients progressing after treatment with hormonal therapies have intrinsic resistance to systemic therapy. The two-thirds which respond, suffer visible side effects from drug toxicity and rapidly acquire resistance to therapy. At present, clinical data to delineate properly the differences between the taxanes is still missing, i.e., one can identify and quantify differences in computational metrics from research point of view, but the clinical angle is limited to cabazitaxel being a more potent version of docetaxel. Novel tubulin inhibitors will be introduced in the clinic and used at earlier stages of disease progression, and once this is done, better links between clinical outcomes and research will be established. Overall, the treatment of advanced PC is hampered by the lack of known molecular markers for reliable targeted therapies. In this contribution, we have proposed methods to anticipate drug resistance in patient cells *ex vivo* and inform therapy. We have also discussed an approach for sensitizing resistant prostate tumors by targeting the tubulin transcriptome. Further, checkpoint blockade inhibitors provide responses of unprecedented durability for a fraction of patients with otherwise treatment-refractory solid tumors. A tumor with high patient tumor-infiltrating lymphocytes (TILs) could have hundreds or thousands of T cells and other immune cells that are all tumor-specific (Zorko et al., 2024). However, the majority of patients experience primary resistance due to low TIL cell infiltration. In PC, a 40 ml peripheral blood draw would not contain a sufficient number of tumor-specific TIL cells and to make immunotherapy feasible, TILs can be isolated, cultured (starting by plating 10 tumor TIL cells per well), and expanded *ex vivo*. Next, a chemical screen can identify small molecule enhancers of TIL cell response to patient-derived tumor organoids. Small molecules can alter the immunogenicity of tumor cells and promote tumor-specific T cell and natural killer cell activation (Mitchison, 2021), and drug candidates can be identified via an unbiased cell-based screen in a tumor organoid and immune cell co-culture format utilizing patient-derived tumor and matched blood samples. T cell and natural killer cell activation can be assayed by high-throughput measurements of cytokine production and quantitative cell biological profiling (Matov, 2024f).

No one has been cured of metastatic PC. Patient tumors are treated with drugs that lower the androgen levels, which induces Alzheimer’s disease (Zhang et al., 2024). Even if approved only in the metastatic setting, androgen-targeting therapy is used off-label in some hospitals to treat primary tumors as well. Androgen deprivation initially lowers patient PSA levels and keeps the tumors at bay for a few years. The disease, unfortunately, once treated with androgen inhibitors, always returns in a very aggressive form. Slow-proliferating heterogeneous prostate tumors then evolve to a fast-proliferating clonogenic metastatic disease for which there are very limited treatment options. Prostate tumor cells (unlike prostate cells derived from normal tissues) cannot be easily cultured *ex vivo* or transplanted in PDX models and that indicates the existence of a specific for the human body microenvironment in PC, which is poorly understood. The fact, however, that more and more clinical studies recommend the utilization of tubulin inhibitors earlier in disease progression, including as neo-adjuvant therapy (Sumiyoshi et al., 2024), suggests an interplay between AR and MT regulation. GSK3 is a well-established component of the Wnt signaling pathway and one of the most prominent regulators of MT homeostasis (Frame and Cohen, 2001). This kinase sits in the signal transduction and phosphorylation cascade one level above the immediate MT-associated proteins and regulates their behavior (Hajka et al., 2021). GSK3 inhibitors are currently being tested for over 20 clinical indications from neurodegeneration to a variety of cancers (Duda et al., 2020; Wood, 2022). Perhaps targeting such genes, which are located at an earlier stage of signal transduction than the transcription factor AR, could bring the PSA levels to normal values without triggering a cascade of long-term side effects. In addition, different GSK3 isoforms are elevated in early (GSK3α) and advanced disease (GSK3β) (Duda et al., 2020), providing a rationale for differential targeting and giving hope that Gleason Score 3 tumors can be cured. Low grade tumor cells maintain their progenitor cell attributes (Lukacs et al., 2008) and it is conceivable that lowering the levels of GSK3α (Ma et al., 2017) may revert the differentiation processes. Perhaps MT dynamics can offer a functional assay to screen putative compounds that correct aberrant MT homeostasis in disease (Matov, 2024d), such as the increase in MT polymerization rates during interphase, which likely shortens interphase and accelerates cell cycle progression and mitotic entry in a subset of metastatic PC cells. These agents might be inhibitors of a variety of kinases (Kannaiyan and Mahadevan, 2018) and MT-associated proteins (Fig. 11) (Čermák et al., 2020), or novel compounds which act on the DNA binding (Pang et al., 2022) and transactivation (Zhu et al., 2022) domains of AR.

**Figure 11.**
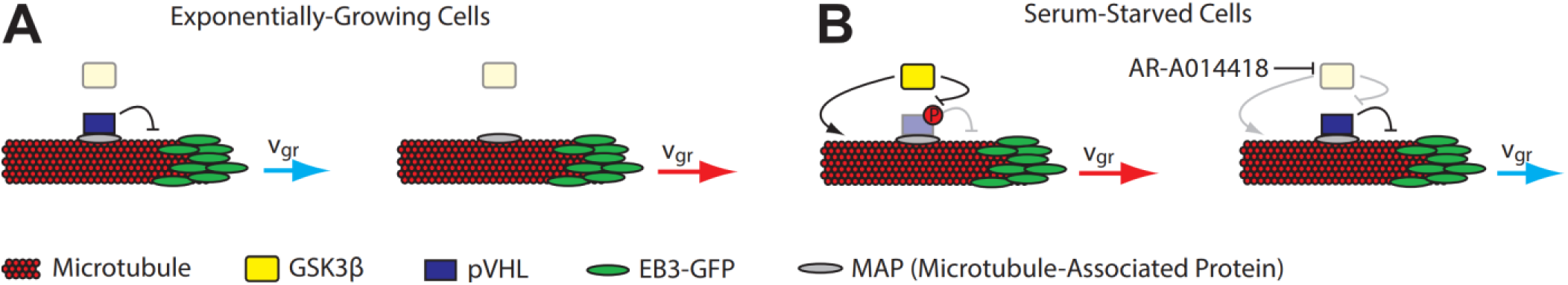
Loss of pVHL in exponentially-growing cells and GSK3β activity in quiescent cells lead to an increase in MT turnover rates. pVHL attenuates MT dynamics by lowering the rates (blue arrows) of tubulin turnover both at growing and shrinking MT tips, and by inhibition of MT catastrophe events likely because pVHL30 reduces the intrinsic GTPase activity (Thoma et al., 2010). These functions of pVHL contribute to the changes of MT dynamics when cells are serum-starved and are negatively regulated by GSK3β. (A) In exponentially-growing cells with a functioning tumor suppressor protein pVHL, the loss of pVHL leads to an increase in the rates of MT polymerization (red arrow). (B) In serum-starved (quiescent) cells, GSK3β phosphorylation of pVHL leads to fast tubulin turnover (red arrow). The inhibition of GSK3 restores pVHL function and baseline MT dynamics. MT-associated proteins, like CLASP2, regulate MT dynamics as well.

When a drug regimen is selected, the selection of drug dose and deducing its effects (Zasadil et al., 2014), depending on the dose administered, is a daunting task. During treatment with tubulin inhibitors, for instance, different MT-associated proteins are activated depending on the drug dose administered, and consequently, different resistance mechanisms may be triggered (Matov, 2024d; Matov, 2024e). The effects of MT inhibitors change nonlinearly with a dose increase. While a high dose of nocodazole depolymerizes MTs, a low dose, which could lead to no side effects, increases polymerization rates (Vid. 6) (Matov, 2024d; Thoma et al., 2010) and that would induce different medical effects. While a high dose of paclitaxel stabilizes MTs, a low dose similarly increases MT polymerization rates, which can, counterproductively, assist cancer cells in progressing through mitosis (Ertych et al., 2014; Matov, 2024c; Matov, 2024d).

During pathogenesis, or as a result of drug treatment in disease (Gonçalves et al., 2001), cells change their intracellular organization (Galletti et al., 2014; Matov, 2024d), rearrange their internal components as they grow, divide, and adapt mechanically to a hostile environment. These functions depend on the controlled polymer turnover of protein filaments (the cytoskeleton and FAs), which provide the cell shape (Matov, 2024a) and its capacity for directed movement (Lauffenburger and Horwitz, 1996) and, thus, determine tumor aggressiveness and sensitivity to drug treatment. In this context, specialized image analysis approaches (Matov, 2024f) for the quantification of the physiological intercellular protein dynamics (Matov, 2024f) in patient-derived cells might become an indispensable tool for drug development as well as the selection of optimal drug regimens (Matov, 2025a) and dose selection in both organ-confined and metastatic PC.

## MATERIALS AND METHODS

### Coverslip Processing

Peripheral blood previously drawn from mCRPC patients (the samples were de-identified) receiving docetaxel-based first line chemotherapy was thawed and processed using ficolling (Ficoll-Paque®) to separate cells in the buffy coat from the blood. The cells were isolated by centrifugation, fixed in phenol, and blocked on 8 mm coverslips. In brief, the ficolling method, through centrifugation, isolates peripheral blood mononuclear cells and plasma from granulocytes and erythrocytes. The mononuclear layer contains lymphocytes, monocytes, and CTCs. These cells are plated onto an 8 mm coverslip and subsequently stained for biomarkers (PSMA, CD45, and DAPI). PSMA labeling was done using ^177^Lu - J591, anti-PSMA monoclonal antibody J591 (Matov and Modiri, 2024).

### Cell Culture

Primary and retroperitoneal lymph node metastatic prostate tumor and sternum metastatic rectal tumor tissues were dissociated to single cells using modified protocols from the Witte lab. Organoids were seeded as single cells in three 30 µL Matrigel drops in 6-well plates. Organoid medium was prepared according to modified protocols from the Clevers lab and the Chen lab.

To obtain a single cell suspension, tissues were mechanically disrupted and digested with 5 mg/ml collagenase in advanced DMEM/F12 tissue culture medium for several hours (between 2 and 12 hours, depending on the biopsy/resection performed). If this step yielded too much contamination with non-epithelial cells, for instance during processing of primary prostate tumors, the protocol incorporated additional washes and red blood cell lysis (Goldstein et al., 2011). Single cells were then counted using a hemocytometer to estimate the number of tumor cells in the sample, and seeded in growth factor-reduced Matrigel drops overlaid with prostate cancer medium (Gao et al., 2014). With radical prostatectomy specimens, we had good success with seeding three thousand cells per 30 μl Matrigel drop, but for metastatic samples organoid seeding could reliably be accomplished with significantly less cells, in the hundreds. To derive organoids from patient CTCs, liquid biopsy samples of 40 ml peripheral blood would be collected, processed, and plated in a Matrigel-Collagen-Fibronectin matrix to form organoids similarly to the metastatic breast cancer organoids we cultured from mouse CTCs (Fig. 12).

**Figure 12.**
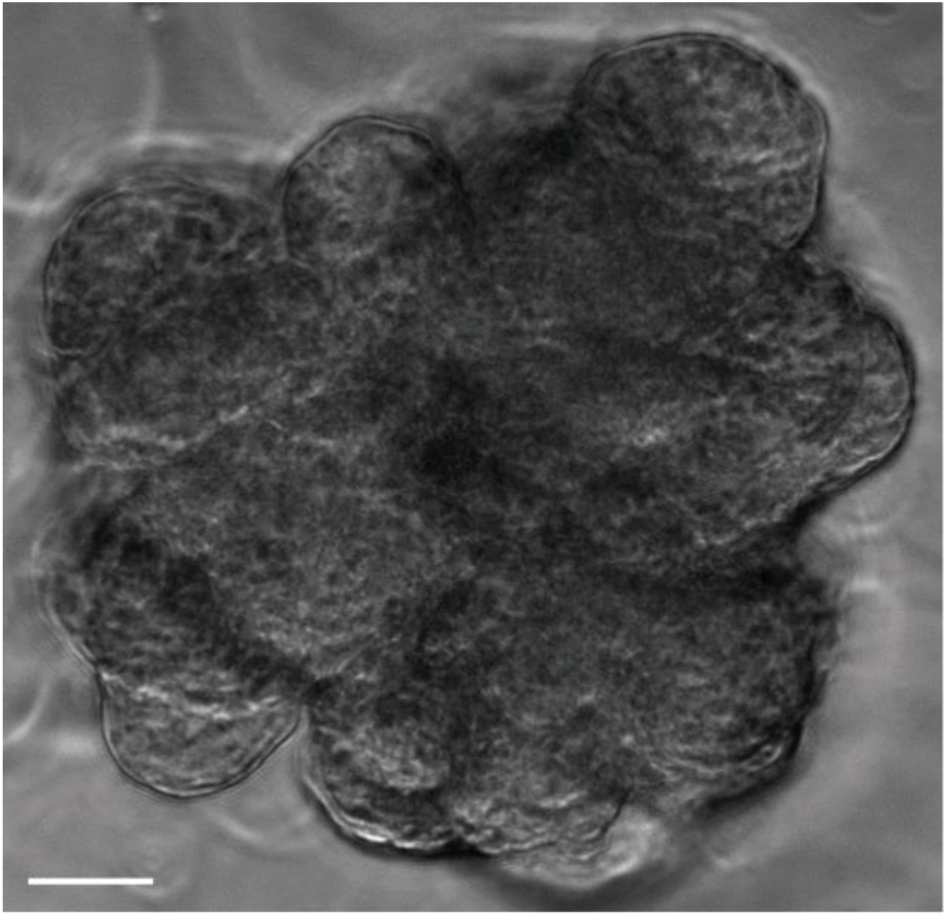
Phase contrast microscopy image of a breast cancer organoid. Organoids derived from CTCs from a mouse xenograft model of invasive ductal breast carcinoma. Scale bar equals 30 µm.

To transduce organoids, we modified protocols from the Clevers lab to adapt to the specifics of prostate organoid culture (such as the significant differences in proliferation rates in comparison to colon and rectal organoids). We found out empirically that cells in mid-size organoids (60-100 µm in diameter) infect at much better rates than trypsinized single organoid cells. These were the steps we followed to express EB1ΔC-2xEGFP in organoids: (1) Add Dispase (1 mg/ml) to each well to dissolve the Matrigel at room temperature for 1 hour. (2) Spin down (at 1,000 rpm for 4 minutes) and mix organoids with 10 µl of viral particles (enough for 1 well with three Matrigel drops of 30-40 µl with organoids containing 1-2 million cells) with Y27632 ROCK inhibitor and Polybrene (1:1,000) for 30 minutes. (3) Spin the organoids with viral particles for 1 hour at 600g. (4) Leave the organoids for a 6-hour incubation. (5) Spin down and plate in Matrigel. (6) 1 hour later, add medium. We used blasticidin (1:20,000) for only one round of medium (3 days) because an increase of the density of labeled cells in the organoids reduced our ability to image MT tips with good contrast.

### Live-Cell MT Tracking

M12 cells were transfected with plasmid encoding EB1ΔC-2xEGFP (Piehl and Cassimeris, 2003) and EB1 comets in live cells were imaged by spinning-disc confocal microscopy using a 100x magnification oil immersion 1.49 NA objective for all cultured cells as previously described (Gierke and Wittmann, 2012) and tracked using our image analysis algorithm (Matov et al., 2010). EB1 transiently binds to growing MT plus ends (Akhmanova and Steinmetz, 2008), generating a punctate pattern of EB1ΔC-2xEGFP comets throughout the cell. The exponential decay of available binding sites results in the characteristic comet-like fluorescence profiles of EGFP-tagged end-binding proteins (“comets”). MDA-MB-231 cells were transiently transfected with EB1ΔC-2xEGFP and treated with vehicle (DMSO) or treated with 100 nM paclitaxel in for 2 hours. Paclitaxel (in intravenous solution) was a gift from Linda Vahdat. To test for statistically significant shifts in the distributions of MT growth rates obtained following the tracking of 195,970 EB1ΔC-2xEGFP comets, we performed per-MT track and per-cell analysis, and compared the average EB1 speeds for 14 DMSO-treated and 14 paclitaxel-treated cells (see Statistical Analysis). Clear cell renal cell carcinoma (ccRCC) cells were cultured and imaged as previously described (Thoma et al., 2010). Receptor triple negative breast cancer (MDA-MB-231) cells were cultured and imaged as previously described (Matov, 2024d).

### Microscopy Imaging

The type of microscopy used to image fixed cells was spinning-disc confocal microscopy with a 10x magnification 0.3 NA objective. The acquisition was accomplished using an ORCA-Flash 4.0 (6.5 µm/pixel) camera. We imaged EB1ΔC-2xEGFP-expressing organoids (Koo et al., 2013) by time-lapse spinning disk confocal microscopy using a long working distance 60x magnification water immersion 1.45 NA objective. We acquired images at half a second intervals for a minute to collect datasets without photobleaching (Gierke and Wittmann, 2012).

### Image Analysis

All image analysis programs for drug-induced tubulin bundling, EB1/3 comet detection, MT dynamics, and graphical representation of the results were developed in Matlab and C/C++. The computer code is available for download at: https://github.com/amatov/ClusterTrackTubuline and https://github.com/amatov/ResistanceBiomarkerAnalysis.

### Statistical Analysis

Statistical tests were performed in Matlab. We perform Kolmogorov-Smirnoff statistics (Kolmogorov, 1933) to compare high order moments of the underlying MT parameter per track distributions and permutation *t*-test to extract information on shifts in median values of polymerization parameters over a population of cells. In brief, for each condition, multiple videos containing one or two cells were compared. Before merging the data for further statistical analysis, we determined the video to video variation in the distributions of MT parameters using the Kolmogorov-Smirnov test (Kolmogorov, 1933). Distributions were subsampled to 400 values to avoid hypersensitivity of the Kolmogorov-Smirnov statistics. Videos passing the consistency test were included in the final datasets. Statistical comparison of MT dynamics between different conditions was accomplished in two steps. First, we examined whether the different conditions would profoundly change the distribution of each parameter, which would indicate an entirely different state of MT regulation between conditions. To test this possibility, we aligned the distributions of compared conditions at their respective per track mean value and then applied the nonparametric Kolmogorov-Smirnov test to determine whether the two distributions are identical. The distributions were randomly subsampled to 400 values. The subsampling was repeated 200 times, and the final p-values were derived by averaging. In a second step, we compared the shifts in parameter mean values between different conditions. To achieve this, we calculated the per cell means for each parameter. Then we tested normality of the distribution of the means and applied a test to define the significance of parameter changes between conditions (mean ± sem of cell to cell variation). This test assumes that the cell to cell variation is the single biggest source of variation in the mean value of a condition. None of the parameters of MT dynamics followed a Gaussian distribution. To determine the significance of differences in the mean values between parameters under different experimental conditions, we applied a non-parametric permutation *t*-test with 400 repetitions (Hesterberg et al., 2005), which compares the bootstrapped distributions of a condition 1 versus condition 2. This test does not make any assumption about the characteristic of the tested distribution and, thus, is appropriate for application to distributions that are far from being normally distributed. In brief, for each condition 400 values are bootstrap-sampled from the data. In agreement with the central limit theorem (Laplace, 1810), the two bootstrapped distributions always follow a Gaussian distribution and, thus, could be analyzed for differences using a regular Student’s *t*-test.

## Data Availability

All data produced in the present study are available upon reasonable request to the authors

## Ethics Declaration

Approval of tissue requests #14-04 and #16-05 to the UCSF Cancer Center Tissue Core and the Genitourinary Oncology Program was given.

## Data Availability Statement

The datasets used and/or analyzed during the current study are available from the corresponding author upon reasonable request.

## Conflicts of Interest

The author declares no conflicts of interest.

## Funding Statement

No funding was received to assist with the preparation of this manuscript.

## ACKNOWLEDGEMENTS

I thank Benet Pera and Andy Tran for helping me generate the image datasets and Neil Bander for the CTC data shown on Fig. 5. Cladio Thoma performed the RCC-4 and RPE-1 cell culture and imaging, and provided cilia maintenance data. The patient blood samples analyzed were from clinical studies with IRB protocols 0804009740 and 0707009283 at Cornell Medicine. The specialized twin-needle is a gift from Paul Wegener, Epitome Pharmaceuticals. Organoid RNA-sequencing data was shared by Yu Chen. Hao Nguyen performed subcutaneous injection of PC organoid cells into immunodeficient mice. Johan de Rooij derived the breast cancer organoid from mouse CTCs. I am grateful to the Genitourinary Tissue Utilization committee and the Genitourinary and Prostate SPORE Tissue Cores at the UCSF Cancer Center for the approval of my tissue requests #14-04 and #16-5 and the Stand Up To Cancer / Prostate Cancer Foundation (SU2C/PCF) West Coast Dream Team (WCDT).

## SUPPLEMENTARY MATERIALS

Video 1 – PC organoid (MSK-PCa5) derived from CTCs with labeled MT tips and overlaid EB1 comet detections.avi. The organoid was established at MSKCC from CTCs of a mCRPC patient with a high CTC count (>100 cells per 8 ml of blood) (Gao et al., 2014). MT tips were labeled, imaged, and computationally detected (green dots in the video; the cell diameter is 10 μm) as visible EB1ΔC-2xEGFP comets (Matov et al., 2010). https://vimeo.com/999622700/4dec3ccc4b

Video 2 – RCC-4 VHL^-/-^ nocodazole 40nM EB3ΔC-2xEGFP comet detections. Detection of visible EB3ΔC-2xEGFP comets (Matov et al., 2010) 20 seconds after the addition of nocodazole. https://vimeo.com/1002865709/1488944d3d

Video 3 – RPE-1 ns-control EB3ΔC-2xEGFP (Thoma et al., 2010) with overlaid clusters of collinear MT tracks (Matov et al., 2010). Visible stages of MT growth are shown in white color, forward gaps (stages of MT pausing) are shown in green color, and backward gaps (stages of MT shrinking) are shown in red color. https://vimeo.com/1002857919/26b19261d0

Video 4 – RPE-1 *shVHL* EB3ΔC-2xEGFP with overlaid clusters of collinear MT tracks (Matov et al., 2010). Visible stages of MT growth are shown in white color, forward gaps (stages of MT pausing) are shown in green color, and backward gaps (stages of MT shrinking) are shown in red color. https://vimeo.com/1002862159/ab7d67e0b2

Video 5 – RPE-1 ns-control CLIP170-2xEGFP comet detections (Matov et al., 2010). https://vimeo.com/1002880407/8d80e5393a

Video 6 – RCC-4 pVHL30 nocodazole 40nM EB3ΔC-2xEGFP comet detections. Detection of visible EB3ΔC-2xEGFP comets (Matov et al., 2010) 20 seconds after the addition of nocodazole. https://vimeo.com/1002869396/48defc570c

The movies have a length of 12.5 seconds. Original time-lapse sequences consist of 125 frames acquired with a frame rate of 0.5 seconds. The replay rate of the movies is 10 frames per second, i.e., a five-fold acceleration.

**Figure S1.**
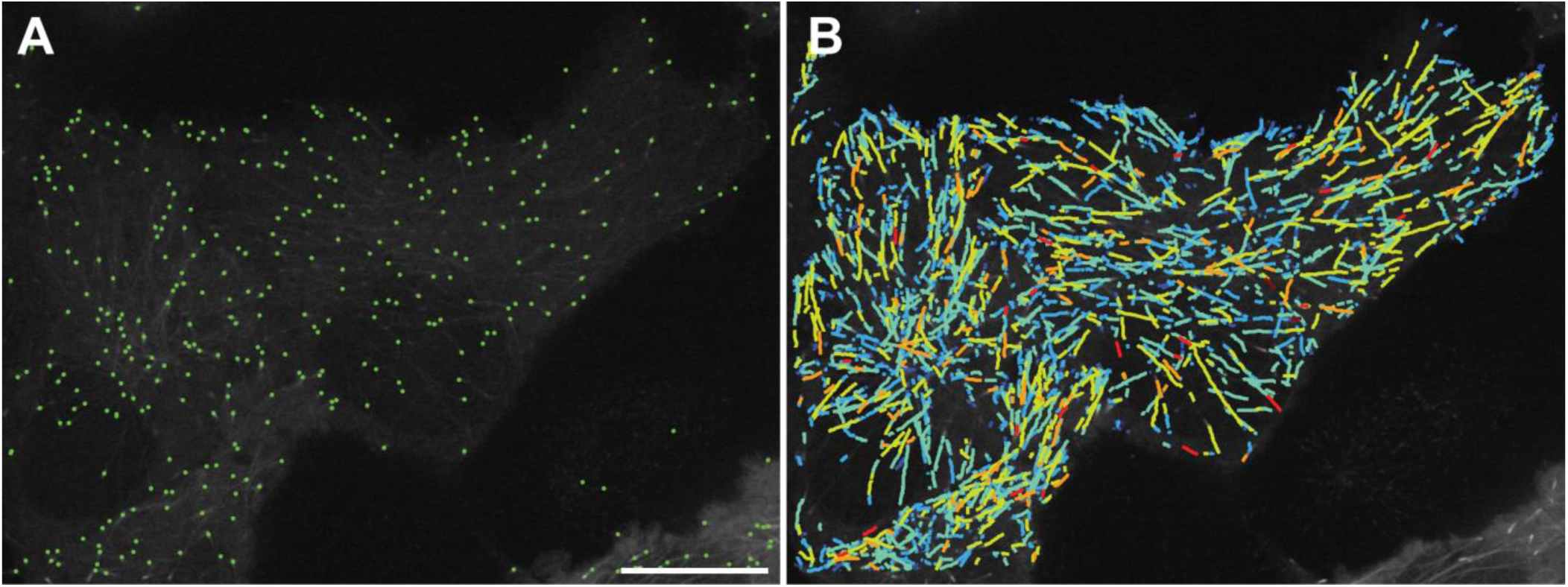
Tracking of EB1 comets in M12 parental androgen-independent cell line. (A) Detection of growing MT tips (Matov et al., 2010) indicates high density of polymerizing MTs and EB1 comets, whose center of mass is displayed with a green dot. (B) MT tips are labeled with GFP, imaged for a minute (with an acquisition rate of two images per second), and EB1 comets are computationally tracked (Yang et al., 2005). The average MT growth rates and EB1 speeds in parental M12 cells not expressing AR is 17.3 μm/min, which is reduced with about 1 μm/min when AR-wt is expressed and increased with over 7 μm/min when the taxane-resistant Arv7 variant is expressed. The color-coding represents EB1 speeds and colder colors correspond to lower speeds, and warmer colors correspond to faster speeds. The scale bar equals 5 µm.

**Figure S2.**
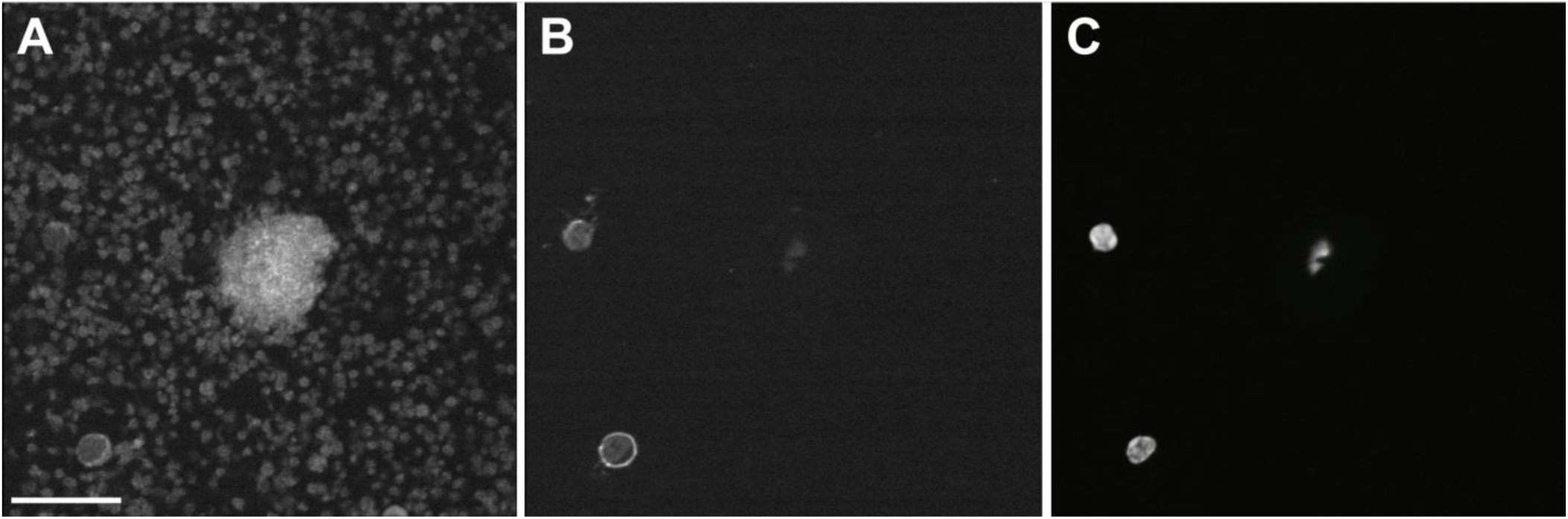
CTC with diameter larger than that of a normal prostate epithelial cell. Multiplex 10x transmitted light microscopy images of a patient blood sample (Matov and Modiri, 2024). One CTC and two leukocytes are visible in this field of view. (A) PSMA (J591, tumor marker). (B) CD45 (leukocyte marker). (C) DAPI (nuclear marker). The scale bar equals 20 µm.

**Figure S3.**
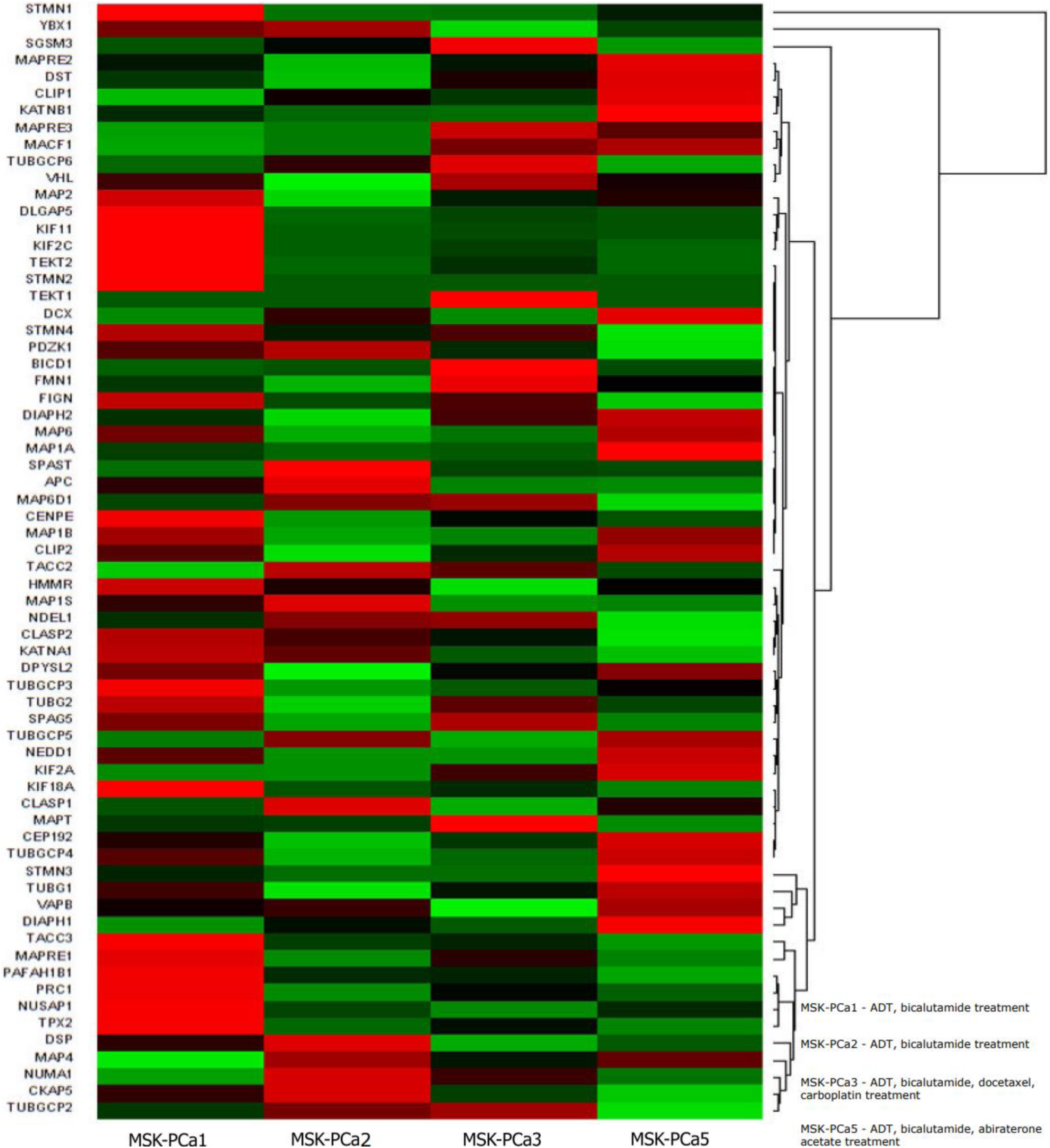
Gene expression signatures of 70 MT regulating genes for MSKCC patient organoids. GSK3 regulates most of the MT regulating genes (Hajka et al., 2021). Hierarchical clustering results indicate that STMN1/Op18 is upregulated in MSK-PCa1, for which we measured attenuated MT polymerization (growth) rates (Matov, 2024d). MT growth rates [µm/min] in cells from organoids PCa1, PCa2, PCa3, and PCa5 are regulated by multiple genes directly, such as chTOG, MAP2, MAP4, CLIP170, and indirectly by other, such as Op18, TPX2 (Lyle et al., 2009a; Lyle et al., 2009b). MT polymerization times [seconds] are reversely correlated with MT catastrophe frequency, i.e., the longer the lifetime of a measured EB1 track is, the less likely it is the MT to switch its state to shrinking. MT depolymerization rates [µm/min] reflect the speed with which MTs remove dimers from their lattice during shrinking. The regulation of stages of MT depolymerization is not well understood. MT depolymerization times [seconds] are reversely correlated with the MT rescue frequency, i.e., the longer a MT is shrinking, the less likely it is to switch its state to growth. Pausing times [seconds] reflect the moments MTs neither grow nor shrink. Our work on the pVHL tumor suppressor protein demonstrated that pausing times are significantly increased when the tumor-suppressor protein is active and reduces the effects of the MT-depolymerizing drug nocodazole (Thoma et al., 2010).

**Figure S4.**
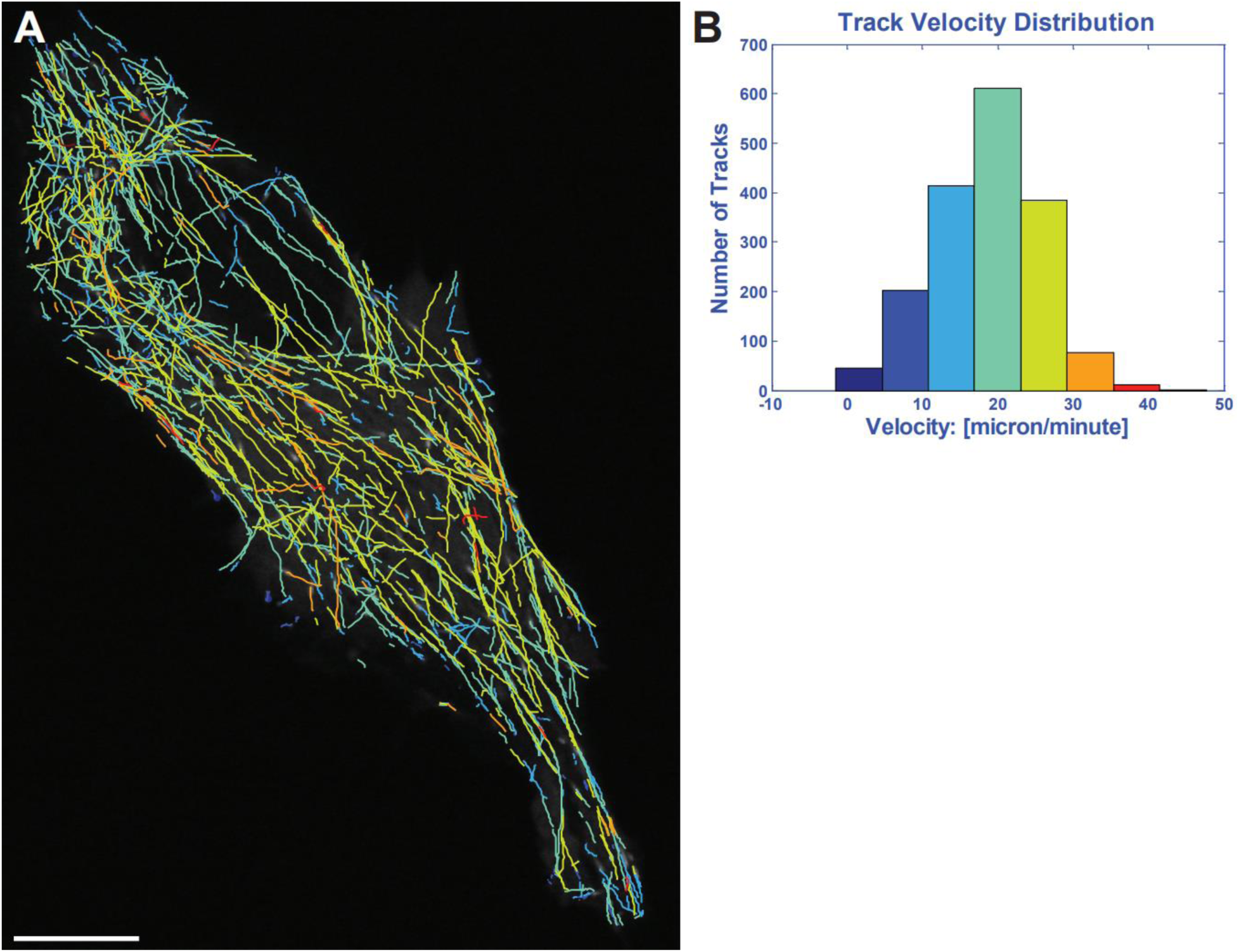
Tracking of EB1 comets in MCF10A normal breast epithelium cell line. (A) MT tips are labeled with GFP, imaged for a minute (with an acquisition rate of two images per second), and EB1 comets are computationally tracked (Yang et al., 2005). The color-coding represents EB1 speeds and colder colors correspond to lower speeds, and warmer colors correspond to faster speeds. The scale bar equals 5 µm. Histogram of growth velocities in µm/min is shown on (B) and indicates a normal distribution and suggests that the EB1 speeds (with a median value of about 20 µm/min and an average of 18.7 µm/min), resulting from a faster MT polymerization in normal breast epithelium, are about 30% to 70% faster (79,560 EB1 comets in MCF10A cells analyzed) in comparison to breast cancer cells (see Fig. 6 and (Matov, 2024d)). Nicotinamide adenine dinucleotide (NAD+) is an endogenous small molecule that has effects on diverse processes, including obesity, lifespan, and cancer, which can increase MT polymerization rates (Harkcom et al., 2014) and restore normal MT regulation and homeostasis.

## Notes

### Competing Interest Statement

The authors have declared no competing interest.

### Funding Statement

This study did not receive any funding

### Author Declarations

The patient blood samples analyzed were from clinical studies with IRB protocols 0804009740 and 0707009283 at Cornell Medicine. I am grateful to the Genitourinary Tissue Utilization committee and the Genitourinary and Prostate SPORE Tissue Cores at the UCSF Cancer Center for the approval of my tissue requests #14-04 and #16-5 and the Stand Up To Cancer / Prostate Cancer Foundation (SU2C/PCF) West Coast Dream Team (WCDT).

### Summary of Updates

Made formatting changes to the manuscript.

